# Associations between spatial distribution of immune cell subsets and clinical outcomes in patients with advanced melanoma treated with immune checkpoint inhibitors: results from the PUMA challenge

**DOI:** 10.64898/2026.03.09.26347935

**Authors:** Mark Schuiveling, Hong Liu, Daniel Eek, Martina Hanusová, Isabella A.J. van Duin, Laurens S. ter Maat, Janneke C. van der Weerd, Franchette van den Berkmortel, Christian U. Blank, Gerben E. Breimer, Femke H. Burgers, Marye Boers-Sonderen, Alfons J.M. van den Eertwegh, Jan Willem B. de Groot, John B.A.G. Haanen, Geke A.P. Hospers, Ellen Kapiteijn, Djura Piersma, Lieke Simkens, Hans M. Westgeest, Anne M.R. Schrader, Paul J. van Diest, Jiaqi Lv, Yijie Zhu, Carmen Guadalupe Colin Tenorio, Brinder Singh Chohan, Mark Eastwood, Shan E Ahmed Raza, Nima Torbati, Anastasia Meshcheryakova, Diana Mechtcheriakova, Amirreza Mahbod, Daniel Adams, Adrian Galdran, Josien P.W. Pluim, Willeke A.M. Blokx, Karijn P.M. Suijkerbuijk, Mitko Veta

**Author notes:** Corresponding author: Mark Schuiveling, MD, Department of Medical Oncology, University Medical Center Utrecht, Utrecht University, Heidelberglaan 100, 3584 CX, Utrecht, the Netherlands. These authors contributed equally to this work (Mark Schuiveling, Hong Liu). These authors contributed equally to this work (Willeke A.M. Blokx, Karijn P.M. Suijkerbuijk, Mitko Veta).

## Abstract

Patients with advanced melanoma are treated with immune checkpoint inhibitors (ICIs), yet less than 50% of patients achieve a durable response while all patients are exposed to the risk of severe side effects. Tumor-infiltrating lymphocytes (TILs) in pathology images are associated with ICI outcomes, but manual assessment is subjective. In addition, the predictive value of other immune cell subsets, including plasma cells, neutrophils, histiocytes, and melanophages, remains unclear. We organized the Panoptic segmentation of nUclei and tissue in advanced MelanomA (PUMA) challenge to evaluate whether the spatial localization of TILs and other immune cell subsets on melanoma H&E slides collected before start of treatment was associated with treatment outcomes. Algorithm performance was evaluated on a hidden test set, after which top-ranked algorithms were applied to pre-treatment metastatic whole-slide images from a large, multicenter cohort of patients with advanced melanoma treated with first-line ICIs (*n*=1102). Automatically quantified tissue features and immune cell subsets were then associated with clinical outcomes.

Top-performing algorithms improved detection of immune cell subsets, although accuracy for rare classes remained limited. Across challenge participants, TIL density showed the most consistent association with treatment response and survival. Associations for stromal TILs were weaker, while plasma cells, histiocytes, melanophages, neutrophils, necrosis and blood vessels did not show independent associations with outcomes. Overall, the results from the PUMA challenge improved the state of the art of immune cell detection in melanoma histopathology and show that intra-tumoral lymphocytes are the immune cell subset most consistently associated with treatment response and survival.

**Highlights:**

- We organized the first melanoma-specific tissue and nuclei segmentation competition
- Winning algorithms were applied to 1102 whole-slide images for biomarker analysis
- Intra-tumoral TILs were associated with response to immune checkpoint inhibitors
- Other immune cell subsets showed no independent association with treatment outcomes
- Tissue segmentation on WSIs was limited by low heterogeneity in training data.

**Graphical abstract:** 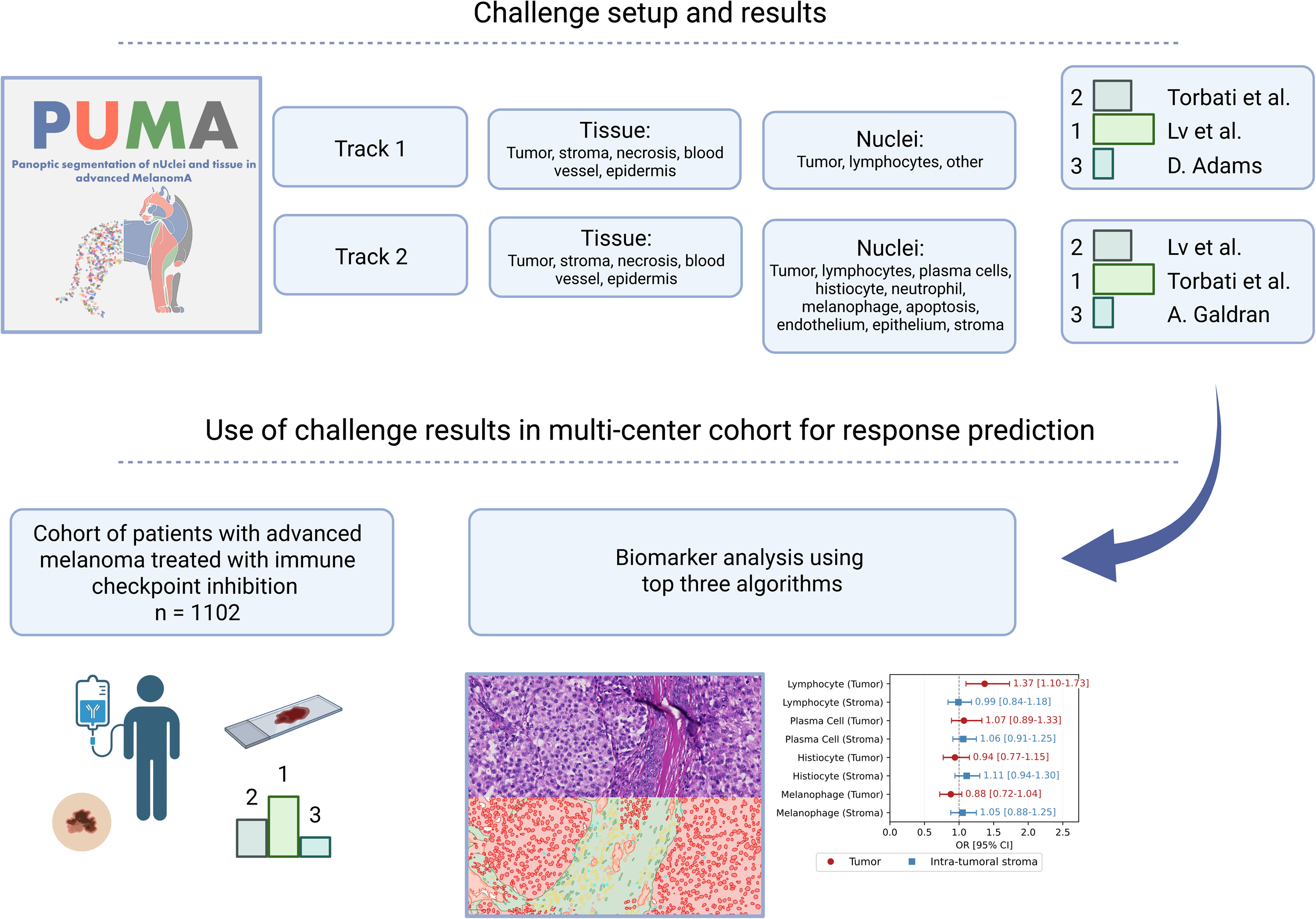

## 1. Introduction

Melanoma is an aggressive skin cancer with an increasing incidence (Rahib et al., 2021). While early-stage melanoma can be cured through surgical excision, advanced melanoma is treated with systemic therapy, mostly consisting of immune checkpoint inhibitors (ICIs). ICIs restore antitumor immune activity by blocking inhibitory receptors like PD-1 and CTLA-4, resulting in durable responses and potential cures in up to half of patients (Wolchok et al., 2022). However, it also results in severe and sometimes irreversible toxicity affecting up to 60% of patients treated with combination therapy, underscoring the need for reliable predictive biomarkers (Suijkerbuijk et al., 2024).

We previously identified manually scored tumor-infiltrating lymphocytes, TILs, as a potential biomarker of response (Duin et al., 2024). Manual assessment, unfortunately, also showed considerable interobserver variability, limiting clinical applicability. Automated deep learning–based detection offers a more consistent alternative, yet existing TIL detection models performed poorly in melanoma due to its ability to mimic other cell types (Mark Schuiveling et al., 2025a). We, therefore, developed an open-source, melanoma-specific dataset with detailed nuclei annotations (Mark Schuiveling et al., 2025a) with which we retrained an existing TIL detection model called Hover-Next (Baumann et al., 2024). With this approach we demonstrated that AI-detected TILs correlate stronger with treatment outcomes than manually scored TILs (Mark Schuiveling et al., 2025b).

In addition to TILs, the dataset includes annotations for visually distinguishable immune cell types such as plasma cells, histiocytes, melanophages, and neutrophils. This was motivated by studies in other tumor types showing that several of these subsets, identified through immunohistochemistry or RNA sequencing, independently associate with treatment outcomes (Patil et al., 2022; Zhang et al., 2024). However, our previous research demonstrated that it is not possible with high accuracy to detect these cell types using of the shelve nuclei detection models (Mark Schuiveling et al., 2025a).

We hypothesized that the detection of there rare classes could be improved by the integration of tissue and nuclei detection as this is more in line with the hierarchical approach of analysis that pathologist use to interpret histopathology. This resulted in the inclusion of tissue annotations in the dataset. This also enables spatial assessment of TILs, distinguishing those located within the tumor from those in the tumoral stromal regions, with intratumoral TILs showing the strongest association with treatment outcomes in other tumor types treated with ICIs (Shen et al., 2024).

To test our hypothesis, and to assess the correlation between spatial immune cell subset location and treatment outcomes, we organized the Panoptic segmentation of nUclei and tissue in advanced MelanomA (PUMA) Grand Challenge, hosted on the grand-challenge.org platform. In this competition, participants were invited to develop algorithms for joint segmentation of tissue and nuclei in melanoma histopathology slides. The challenge can be found on https://puma.grand-challenge.org/.

The challenge consisted of two tracks: Track 1 focused on detection of tumor cells, TILs, and other nuclei types along with all tissue classes, aiming to assess whether improved detection of TILs and assessment of stromal or intra-tumoral TILs would better correlate with treatment outcomes. Track 2 expanded the tasks to include additional nuclei types (plasma cells, neutrophils, histiocytes, endothelial cells, stromal cells, apoptotic cells, epithelial cells, and melanophages), again combined with all tissue classes. This track was exploratory, aiming to determine whether other immune cell subsets detectable on H&E slides also hold predictive value.

In this paper, we present the outcomes of the PUMA Grand Challenge, evaluating the performance of the top teams and describing their methodologies. In the second part of the study, we assess the clinical relevance of the resulting models by validating their performance on pre-treatment metastatic melanoma samples from the PREMIUM cohort; a multicenter cohort of patients with treated with first-line ICI therapy.

## 2. Materials and Methods

### 2.1 PUMA challenge setup

#### 2.1.1. PUMA dataset and baseline algorithm

The PUMA dataset consists of 155 primary and 155 metastatic melanoma regions of interest (ROIs), each derived from a separate melanoma case. All cases were digitized in a large melanoma referral center using a Nanozoomer XR C12000–21/–22 (Hamamatsu Photonics; Hamamatsu) at 40× magnification with a resolution of 0.23 µm per pixel. Out of all cases 76 are revisions or consultations originating from other treatment hospitals. ROIs are hematoxylin and eosin (H&E)–stained and include medical expert created and pathologist (certified dermatopathologist W.A.M.B.) verified tissue and nuclei annotations. The dataset is divided into a publicly available training set containing 103 primary and 103 metastatic ROIs (Schuiveling, 2024) under a CC0 license, a preliminary test set of 5 primary and 5 metastatic samples, and a final test set consisting of 47 primary and 47 metastatic samples. This division ensured that all tissue and cell types were sufficiently present in the training, preliminary test and final test set. Only the challenge organizers and the support team of the grand challenge platform had access to the preliminary and final test dataset. The creation of this dataset and the release of the training data was reviewed and approved by the Biobank Research Ethics Committee of UMC Utrecht in accordance with applicable regulations and laws (TCBio 23-270/U-B).

Results from the PUMA challenge are shown for inference on the 94 ROIs of the final hidden test set of the PUMA challenge. We provided a Hover-NeXt model (Baumann et al., 2024) and NN-Unet model (Isensee et al., 2021) as a baseline for the competition, both were optimized on the training dataset (Schuiveling et al., n.d., n.d.). Further information regarding the PUMA dataset curation and annotation, and the baseline methods can be found in the dataset description paper (Mark Schuiveling et al., 2025a).

#### 2.1.2. Challenge performance evaluation

Model performance was evaluated separately for tissue and nuclei segmentation tasks. Tissue segmentation was evaluated with the Dice score. When both the ground truth and the prediction are empty, the Dice score is mathematically undefined. However, standard practice explicitly defines the score as 1 in this case to reflect that the model correctly predicted the absence of the region. This can result in inflated high average (over the evaluation set) Dice scores if a tissue class is present in only a few samples. To accommodate for this, we calculated the micro-average Dice. This is the Dice score for all predictions concatenated along one axis, resulting in a prediction mask of 1,024 × 96,256 pixels. The micro-average Dice is reported with a 95% confidence interval, generated through bootstrapping the sample results (Maier-Hein et al., 2024).

Nuclei segmentation performance was assessed using the *F*_1_ score. Predicted and ground-truth nuclei were matched based on centroid distance, with matches defined within a 15-pixel (3.3 µm) radius, smaller than the average lymphocyte diameter, representing the smallest nuclei in the dataset. When multiple predictions were within this range, the match was assigned based on the highest prediction score (if available) or the shortest centroid distance. Ground-truth nuclei without a match were counted as false negatives and predicted nuclei without a match were counted as false positives. Summed true positives, false positives, and false negatives were used to calculate class specific *F*_1_ *scores,* an *average F*_1_ *score* (the average of class-specific *F*_1_ scores) is used to enable comparison across participants (Maier-Hein et al., 2024).

The overall score for the challenge was the sum of the rank that participants reached for the *average F*_1_ score for nuclei and the micro tissue dice. Micro tissue dice was chosen as this reflects the detection of rare classes such as necrosis and blood vessels. When ranks were equal for participating teams, the mean of the scores was used to define the higher scoring participant. Evaluation code can be found on GitHub. (Schuiveling et al., n.d., n.d.). Model performance was compared with interobserver scores, which were determined on 12 randomly selected regions of interest (ROIs) annotated independently by author M.S. (reviewed by dermatopathologist W.B.) and a second pathologist (author G.B.). No time limit was set for the evaluation dataset processing and the running time of the algorithms was not part of the evaluation. All results were announced publicly.

#### 2.1.3. Challenge participant methodology

Participants used multiple methodological strategies for improvement upon the baseline algorithms. For tissue segmentations, Lv et al. fused features from the Virchow2 foundation model with RGB input in an encoder–decoder architecture to improve detection of rare tissue classes (Lv et al., 2025b), while Torbati et al. used a multistage design with dedicated models for rare tissue classes (Torbati et al., 2025). D. Adams used a U-Net architecture with a ConvNeXt-Base backbone within a joint tissue–nuclei segmentation framework, applying class-weighted sampling during training to address tissue class imbalance.

For nuclei segmentation, participants used a range of approaches. Torbati et al. and D. Adams incorporated tissue context into nuclei prediction, which improved compartment-aware detection and reduced implausible tumor-cell detections within stromal regions. A. Galdran improved test-set performance through balancing strategies, but generalization to the clinical cohort was limited, likely due to sensitivity to staining variation. Lv et al. developed KongNet, a multi-headed architecture with a shared backbone and class-specific heads, aimed at reducing interference between visually similar cell classes (Lv et al., 2025a).

A full description of the methodology and respective references of the challenge participants can be found in the supplementary methodology. Members of the organization of the organizers were not allowed to join the challenge. All participants were required to submit their methods as Docker containers via the Grand Challenge platform which ensured an independent evaluation. The challenge was started on the 10^th^ of October with the release of the training data with a deadline on the 15^th^ of march 2025. Currently the challenge is still open as a rolling challenge for new submissions.

The algorithms of the top three participants in both tracks were validated on a clinical validation cohort. Subsequently, as a reward for achieving a top-three position, these participants were invited to co-author the final results paper. Participants were allowed to also publish their methodology as a separate paper. Participants were not asked to share their code publicly.

### 2.2. Clinical validation of challenge algorithms

#### 2.2.1. Clinical cohort

Patients in the PREMIUM cohort were retrospectively identified from eleven melanoma treatment centers in the Netherlands using high-quality prospectively collected registry data (Jochems et al., 2017). Patients were included if above 18 years of age and treated with first-line anti-PD1 ± anti-CTLA4 for unresectable stage IIIC or stage IV melanoma from January 1, 2016, until the 1^st^ of January 2023. Disease stage was based on the 8^th^ edition of the AJCC staging system (Gershenwald et al., 2017).

#### 2.2.2. Slide selection

Pretreatment metastatic slides were collected from 30 pathology laboratory archives through Palga, the Dutch nationwide pathology databank (Casparie et al., 2007). For each patient, a single representative H&E-stained slide was selected. In cases with multiple specimens, the largest available specimen before therapy initiation was used. Curation was done by four authors (M.S., I.A.J.v.D., L.t.M., and J.v.d.W.) under supervision of two experienced pathologists (W.A.M.B. and P.J.v.D.). All selected slides were scanned with a Nanozoomer XR C12000-21/-22 (Hamamatsu Photonics, Hamamatsu, Shizuoka, Japan) at 40× magnification with a resolution of 0.23 µm per pixel. The anatomic location of the metastatic lesion was derived from the pathology report or assessed visually when possible (in case of presence of pre-existing normal tissue of the organ). Determination of the nature of the specimen type (resection or biopsy) was visually assessed.

#### 2.2.3. Clinical variables and outcome measures

Clinical data included age at treatment initiation, sex, ICI type, World Health Organization (WHO) performance status, BRAF V600E/K mutation status, serum lactate dehydrogenase (LDH) level, AJCC disease stage, and presence of symptomatic brain metastases. We incorporated the variable ‘symptomatic brain metastases’ in the variable ‘stage of disease’: M1D stage was subcategorized either as ‘M1D-non-symptomatic’ or ‘M1D-symptomatic’. LDH levels were categorized as normal, 1-2 times the upper limit of normal (ULN) or twice the upper limit of normal. WHO performance status was categorized as 0, 1, or 2 and above.

Response evaluation was determined by the treating physician and was based on the Response Evaluation Criteria in Solid Tumors, version 1.1, with melanoma-related death before first response assessment classified as progressive disease (Eisenhauer et al., 2009). Primary outcome was objective response, defined as the best overall response (partial or complete response) within 6 months. Secondary outcomes were progression-free survival (PFS) and overall survival (OS).

#### 2.2.4. Challenge validation pipeline

The challenge validation pipeline consisted of several steps. First, the area of interest containing the tumor, tumoral stroma and tumor-associated necrosis was annotated. Annotation of the area of interest was performed manually (by authors J.v.d.W. and M.S., blinded to the outcome) under the supervision of a dermatopathologist (author W.A.M.B.). Second, whole-slide images were tiled into 1024×1024 pixel tiles with a 6.25% overlap, and no color normalization was applied. Third, tiles in which <1% of the area overlapped the manually annotated regions of interest (tumor, tumoral stroma, and tumor-associated necrosis) were filtered out to reduce final processing time.

Tiles were then passed through the participants’ algorithms for inference, which produced tile-level tissue and nuclei segmentation masks. For tissue segmentation, logits from the participants’ models were used to handle ambiguities arising when the same pixel was covered by multiple overlapping tiles. Because the slide background was underrepresented in the PUMA dataset, most models tended to misclassify the white background as tissue. To filter out these irrelevant predictions, a binary background mask was generated for each tile using the H&E-stained tissue-segmentation method of Kleczek et al. (Kleczek et al., 2020) and applied to remove non-tissue regions before producing the final whole-slide tissue-segmentation mask.

For nuclei segmentation, participant models produced either instance segmentations or bounding-box detections. When segmentation outputs were available, overlapping nuclei instances were matched across tiles based on their spatial extent. When detection boxes were provided, non-maximum suppression was used to remove duplicate detections.

Qualitative evaluation of the resulting nuclei and tissue segmentation was done in QuPath (Bankhead et al., 2017).

#### 2.2.5. Feature extraction

For each model, immune cell counts were aggregated within tumor and intra-tumoral stroma compartments. Cell proportions were calculated relative to the total number of nuclei per compartment and standardized to unit standard deviation to facilitate comparison across models.

#### 2.2.6. Statistical analysis

Continuous variables were summarized using median and total range or interquartile range (IQR), and categorical variables with frequencies and percentages.

Correlations were evaluated with Spearman correlation coefficients. Median follow-up was estimated using the reversed Kaplan-Meier method.

Univariable and multivariable analyses were conducted using logistic regression and Cox proportional hazards regression. Model assumptions were visually checked and showed no violations. Statistical analyses were performed using R (version 4.2.2; survival package version 3.5.0).

#### 2.2.7. Ethical review of study

This study was considered not subjected to the medical research involving human subjects act (medical ethical committee reference number WAG/mb/20/004260).

#### 2.2.8. Text and Code Review

ChatGPT Pro 5.2 was used for spelling and grammar checking. All text was manually reviewed and refined by the authors. GitHub Copilot (with auto model selection) assisted in coding, with all code being manually verified before use.

## 3.1 Results

### 3.1. PUMA challenge results

#### 3.1.1. Track 1

In track 1 of the PUMA challenge the goal was to identify all tissue classes and three nuclei classes (tumor, lymphocytes (including lymphocytes and plasma cells) and other cells.

In total 11 teams participated, with 7 teams improving the performance of the baseline model on segmentation of nuclei (Table 1, Supplementary Table 1). Main improvements were observed in the detection of the tumor and the “other” nuclei classes, with *F*_1_ scores increasing from 0.78 [95% CI: 0.75–0.80] to 0.85 [95% CI: 0.83–0.87] and from 0.51 [95% CI: 0.46–0.56] to 0.61 [95% CI: 0.56–0.66], respectively. In contrast, lymphocyte detection showed only a modest improvement, from 0.79 [95% CI: 0.76–0.81] to 0.82 [95% CI: 0.79–0.84].

**Table 1.**
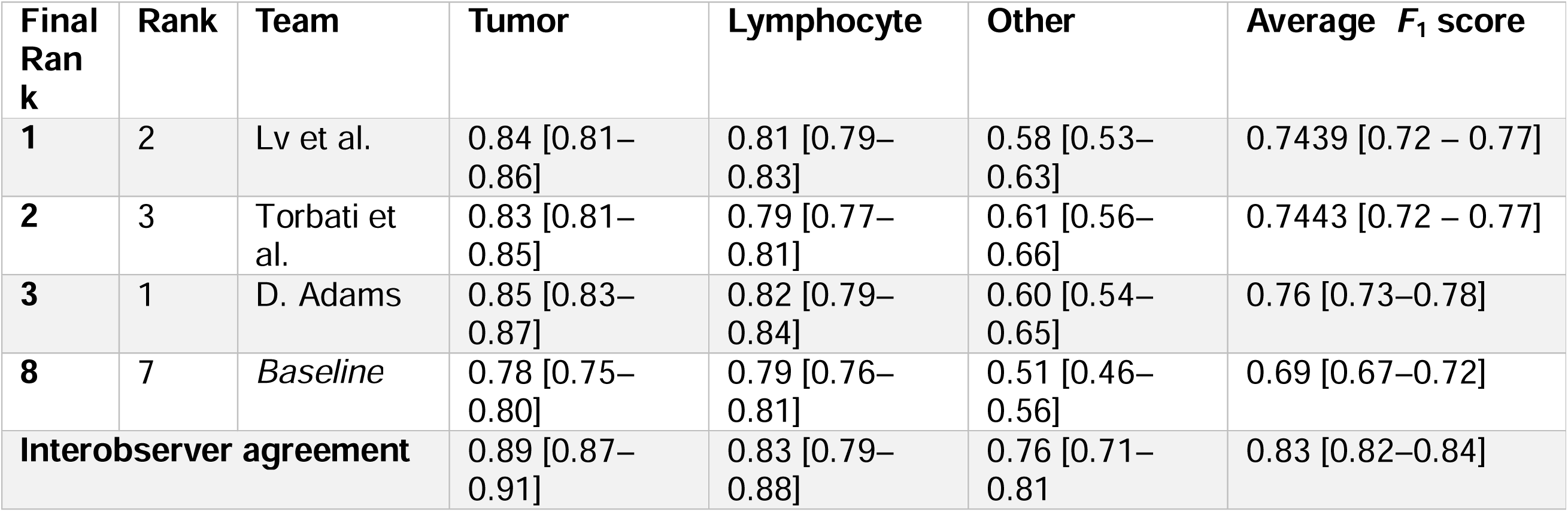
Nuclei segmentation *F*_1_ score of the top three teams and the baseline model in track 1 of the PUMA Grand Challenge. Results are sorted based on the final rank which is the average rank of the tissue and nuclei segmentation task.

For tissue segmentation, the largest improvements were observed in the less common classes, particularly necrosis, with the Dice score increasing from 0.02 [0.00–0.04] to 0.82 [0.02–0.93], and blood vessels, increasing from 0.35 [0.23–0.47] to 0.54 [0.46–0.61]. Performance for more prevalent tissue types also improved, with tumor segmentation increasing from 0.91 [0.88–0.94] to 0.94 [0.92–0.95] and stroma segmentation from 0.79 [0.71–0.84] to 0.84 [0.79–0.88].

**Table 2.**
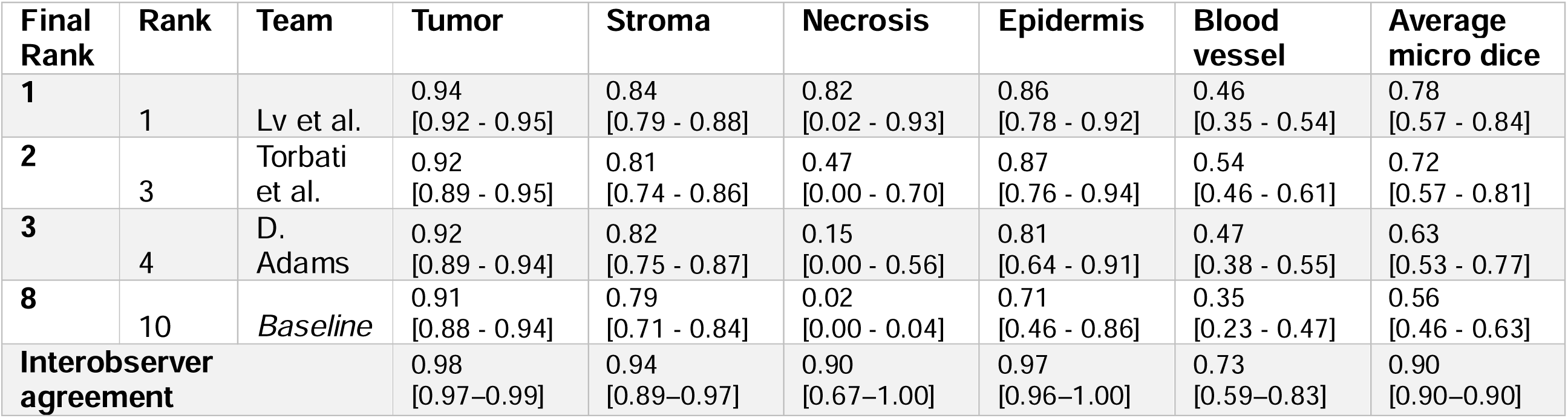
Tissue segmentation dice score of the top three teams and the baseline model in track 1 of the PUMA Grand Challenge. Results are sorted based on the final rank which is the average rank of the tissue and nuclei segmentation task.

| Final Rank | Rank | Team | Tumor | Stroma | Necrosis | Epidermis | Blood vessel | Average micro dice |
| --- | --- | --- | --- | --- | --- | --- | --- | --- |
| 1 | 1 | Lv et al. | 0.94<br>[0.92 - 0.95] | 0.84<br>[0.79 - 0.88] | 0.82<br>[0.02 - 0.93] | 0.86<br>[0.78 - 0.92] | 0.46<br>[0.35 - 0.54] | 0.78<br>[0.57 - 0.84] |
| 2 | 3 | Torbati et al. | 0.92<br>[0.89 - 0.95] | 0.81<br>[0.74 - 0.86] | 0.47<br>[0.00 - 0.70] | 0.87<br>[0.76 - 0.94] | 0.54<br>[0.46 - 0.61] | 0.72<br>[0.57 - 0.81] |
| 3 | 4 | D. Adams | 0.92<br>[0.89 - 0.94] | 0.82<br>[0.75 - 0.87] | 0.15<br>[0.00 - 0.56] | 0.81<br>[0.64 - 0.91] | 0.47<br>[0.38 - 0.55] | 0.63<br>[0.53 - 0.77] |
| 8 | 10 | Baseline | 0.91<br>[0.88 - 0.94] | 0.79<br>[0.71 - 0.84] | 0.02<br>[0.00 - 0.04] | 0.71<br>[0.46 - 0.86] | 0.35<br>[0.23 - 0.47] | 0.56<br>[0.46 - 0.63] |
| Interobserver agreement |  |  | 0.98<br>[0.97–0.99] | 0.94<br>[0.89–0.97] | 0.90<br>[0.67–1.00] | 0.97<br>[0.96–1.00] | 0.73<br>[0.59–0.83] | 0.90<br>[0.90–0.90] |

#### 3.1.2. Track 2

In track 2 of the PUMA challenge, participants were tasked with segmenting all tissue classes and all nuclei subtypes. A total of 12 teams participated, of which 10 outperformed the baseline model on nuclei segmentation (Table 3, Supplementary Table 3).

**Table 3.**
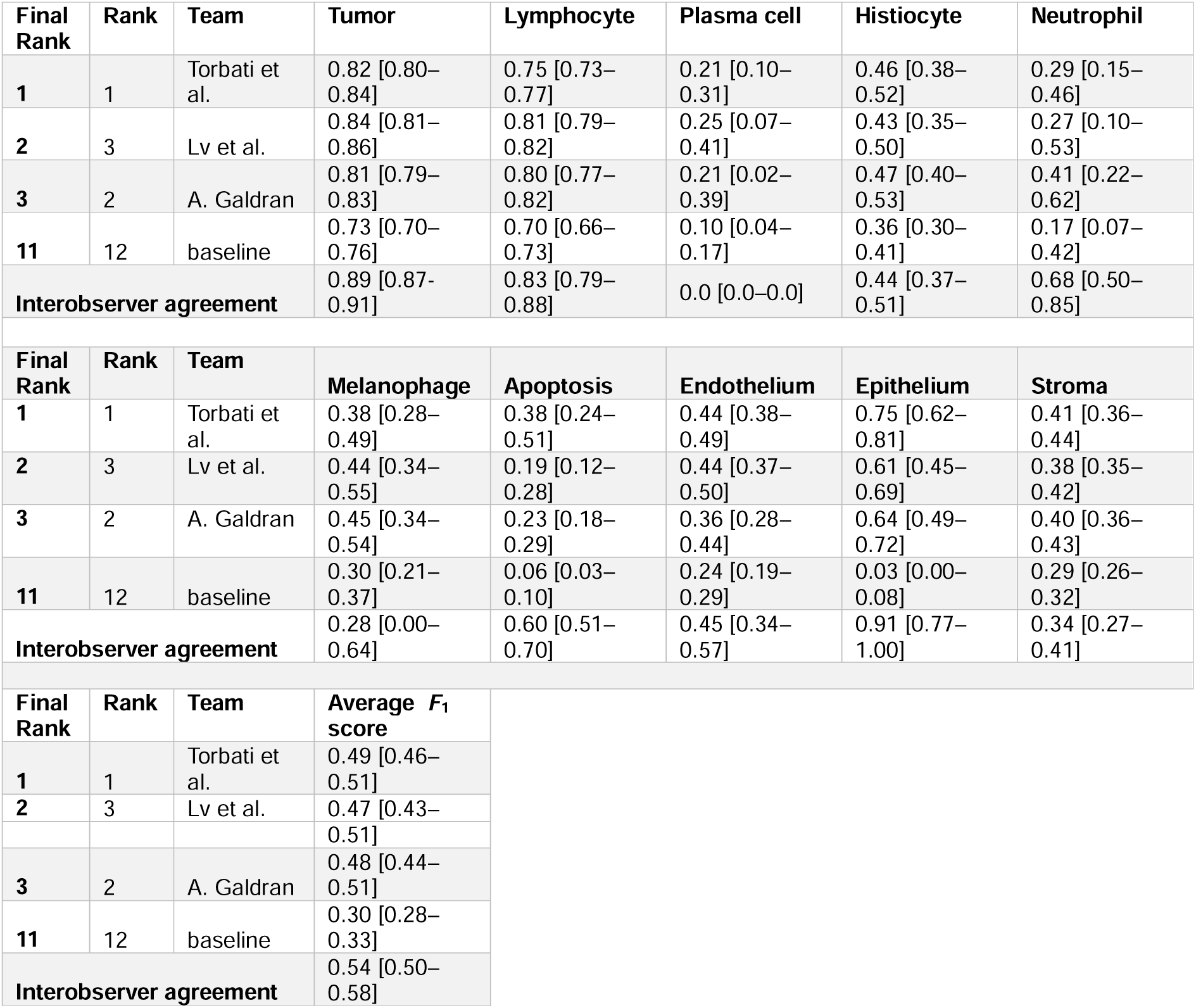
Nuclei segmentation *F*_1_ score of the top three teams and the baseline model in track 2 of the PUMA Grand Challenge. Results are sorted based on the final rank which is the average rank of the tissue and nuclei segmentation task

| Final Rank | Rank | Team | Tumor | Lymphocyte | Plasma cell | Histiocyte | Neutrophil |
| --- | --- | --- | --- | --- | --- | --- | --- |
| 1 | 1 | Torbati et al. | 0.82 [0.80–0.84] | 0.75 [0.73–0.77] | 0.21 [0.10–0.31] | 0.46 [0.38–0.52] | 0.29 [0.15–0.46] |
| 2 | 3 | Lv et al. | 0.84 [0.81–0.86] | 0.81 [0.79–0.82] | 0.25 [0.07–0.41] | 0.43 [0.35–0.50] | 0.27 [0.10–0.53] |
| 3 | 2 | A. Galdran | 0.81 [0.79–0.83] | 0.80 [0.77–0.82] | 0.21 [0.02–0.39] | 0.47 [0.40–0.53] | 0.41 [0.22–0.62] |
| 11 | 12 | baseline | 0.73 [0.70–0.76] | 0.70 [0.66–0.73] | 0.10 [0.04–0.17] | 0.36 [0.30–0.41] | 0.17 [0.07–0.42] |
| Interobserver agreement |  |  | 0.89 [0.87-0.91] | 0.83 [0.79–0.88] | 0.0 [0.0–0.0] | 0.44 [0.37–0.51] | 0.68 [0.50–0.85] |
| Final Rank | Rank | Team | Melanophage | Apoptosis | Endothelium | Epithelium | Stroma |
| 1 | 1 | Torbati et al. | 0.38 [0.28–0.49] | 0.38 [0.24–0.51] | 0.44 [0.38–0.49] | 0.75 [0.62–0.81] | 0.41 [0.36–0.44] |
| 2 | 3 | Lv et al. | 0.44 [0.34–0.55] | 0.19 [0.12–0.28] | 0.44 [0.37–0.50] | 0.61 [0.45–0.69] | 0.38 [0.35–0.42] |
| 3 | 2 | A. Galdran | 0.45 [0.34–0.54] | 0.23 [0.18–0.29] | 0.36 [0.28–0.44] | 0.64 [0.49–0.72] | 0.40 [0.36–0.43] |
| 11 | 12 | baseline | 0.30 [0.21–0.37] | 0.06 [0.03–0.10] | 0.24 [0.19–0.29] | 0.03 [0.00–0.08] | 0.29 [0.26–0.32] |
| Interobserver agreement |  |  | 0.28 [0.00–0.64] | 0.60 [0.51–0.70] | 0.45 [0.34–0.57] | 0.91 [0.77–1.00] | 0.34 [0.27–0.41] |
| Final Rank | Rank | Team | Average $F_1$ score | | | | |
| 1 | 1 | Torbati et al. | 0.49 [0.46–0.51] |  |  |  |  |
| 2 | 3 | Lv et al. | 0.47 [0.43– |  |  |  |  |

|  |  |  |  |
| --- | --- | --- | --- |
|  |  |  | 0.51] |
| **3** | 2 | A. Galdran | 0.48 [0.44–<br>0.51] |
| **11** | 12 | baseline | 0.30 [0.28–<br>0.33] |
| **Interobserver agreement** |  |  | 0.54 [0.50–<br>0.58] |

The best-performing models demonstrated consistent improvements across most nuclei classes, particularly for less common subtypes such as plasma cells, histiocytes, and neutrophils. Despite these improvements, the *F*_1_ scores for these rare classes remained moderate (ranging from 0.21 to 0.47). Nevertheless, the performance was comparable to the interobserver agreement. For neutrophils and apoptotic cells, model performance was lower than interobserver agreement, likely due to their overlapping visual characteristics. The interobserver *F*_1_ score for plasma cells was 0, as only a few plasma cells were present in the subset of regions used for interobserver evaluation.

Performance for the more prevalent nuclei types, including tumor and lymphocytes, was considerably higher, with top *F*_1_ scores around 0.82 and 0.81, respectively. The overall Average *F*_1_ score improved from 0.30 [95% CI: 0.28–0.33] for the baseline model to 0.49 [95% CI: 0.46–0.51] for the best-performing team.

Regarding tissue segmentation, the same pattern as in track 1 was observed, with mainly an increase of Dice scores for less common classes such as necrosis, epidermis and blood vessel (Table 4).

**Table 4.**
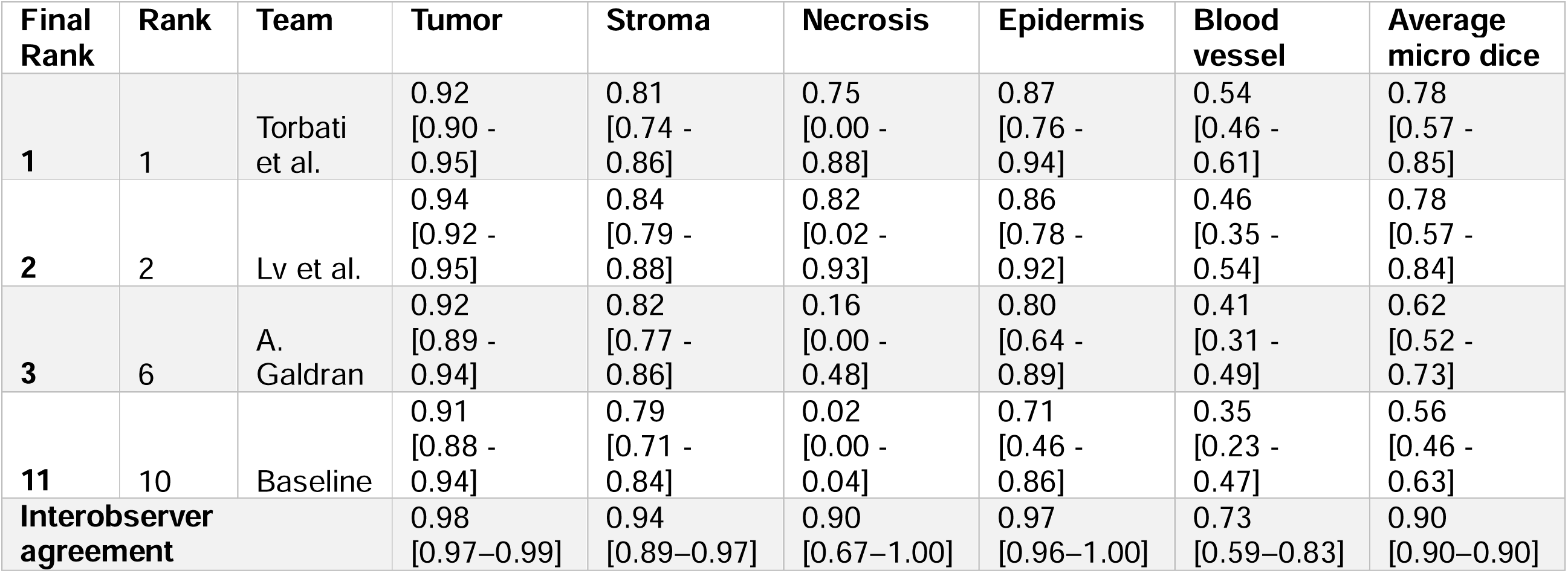
Tissue segmentation performance of the top three teams and the baseline model in track 2 of the PUMA Grand Challenge. Results are sorted based on the final rank which is the average rank of the tissue and nuclei segmentation task.

### 3.2. Clinical validation of challenge algorithms

#### 3.2.1. Patient characteristics and outcomes

The best performing algorithms were evaluated on the multicenter clinical PREMIUM cohort. (Duin et al., 2024; Mark Schuiveling et al., 2025b) From the 1935 patients treated with first-line anti-PD1 +/− anti-CTLA4 therapy, 86 were excluded due to missing treatment response outcome data. Metastatic specimens with viable tumor areas were available for 1177 patients, with 1102 samples successfully processed by all algorithms (flowchart in Supplementary Figure 1). Patient characteristics are shown in Table 5 and compared well to those of excluded patients (Supplementary table 5). Most patients were male, had non-elevated LDH levels, and were above 65 years of age.

**Table 5.**
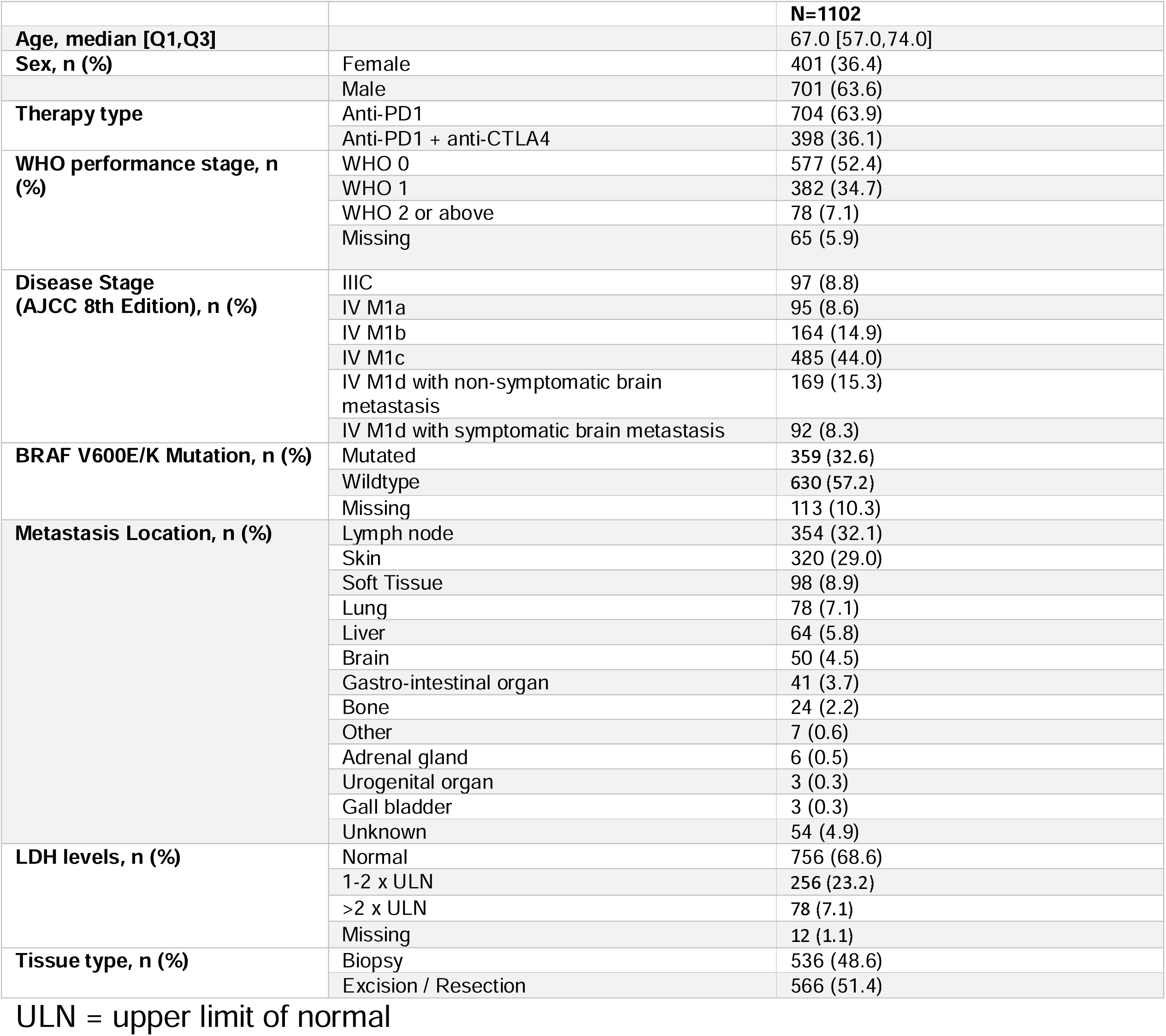
Baseline patient characteristics.

|  |  | **N=1102** |
| --- | --- | --- |
| **Age, median [Q1,Q3]** |  | 67.0 [57.0,74.0] |
| **Sex, n (%)** | Female | 401 (36.4) |
|  | Male | 701 (63.6) |
| **Therapy type** | Anti-PD1 | 704 (63.9) |
|  | Anti-PD1 + anti-CTLA4 | 398 (36.1) |
| **WHO performance stage, n (%)** | WHO 0 | 577 (52.4) |
|  | WHO 1 | 382 (34.7) |
|  | WHO 2 or above | 78 (7.1) |
|  | Missing | 65 (5.9) |
| **Disease Stage (AJCC 8th Edition), n (%)** | IIIC | 97 (8.8) |
|  | IV M1a | 95 (8.6) |
|  | IV M1b | 164 (14.9) |
|  | IV M1c | 485 (44.0) |
|  | IV M1d with non-symptomatic brain metastasis | 169 (15.3) |
|  | IV M1d with symptomatic brain metastasis | 92 (8.3) |
| **BRAF V600E/K Mutation, n (%)** | Mutated | 359 (32.6) |
|  | Wildtype | 630 (57.2) |
|  | Missing | 113 (10.3) |
| **Metastasis Location, n (%)** | Lymph node | 354 (32.1) |
|  | Skin | 320 (29.0) |
|  | Soft Tissue | 98 (8.9) |
|  | Lung | 78 (7.1) |
|  | Liver | 64 (5.8) |
|  | Brain | 50 (4.5) |
|  | Gastro-intestinal organ | 41 (3.7) |
|  | Bone | 24 (2.2) |
|  | Other | 7 (0.6) |
|  | Adrenal gland | 6 (0.5) |
|  | Urogenital organ | 3 (0.3) |
|  | Gall bladder | 3 (0.3) |
|  | Unknown | 54 (4.9) |
| **LDH levels, n (%)** | Normal | 756 (68.6) |
|  | 1-2 x ULN | 256 (23.2) |
|  | >2 x ULN | 78 (7.1) |
|  | Missing | 12 (1.1) |
| **Tissue type, n (%)** | Biopsy | 536 (48.6) |
|  | Excision / Resection | 566 (51.4) |

ULN = upper limit of normal

The median follow-up for patients with a metastatic sample available was 36.3 months [95% CI: 34.37-39.11], with a median progression-free survival (PFS) of 8.8 months [95% CI: 7.87-10.19], and a median overall survival (OS) of 35.9 months [95% CI: 30.62-43.32]. The objective response rate (ORR) to immune checkpoint inhibitors (ICI) was 57.3%.

#### 3.2.2. Tissue segmentation

Tissue segmentation outputs were evaluated for all four teams that ended in the top 3 performing models in both tracks. Across whole-slide images, tumor tissue was the largest proportion of identified tissue, followed by stroma, necrosis, and blood vessels. The model by Torbati et al. identified the highest proportion of necrotic areas, whereas the model by Lv et al. detected relatively more blood vessel tissue (Supplementary Figure 2).

Qualitative assessment of tumor, tumor stroma, and necrosis segmentation demonstrated that, despite the improved segmentation performance in the test set, severe misclassifications farther away from the tumor area persisted. This can be explained by the focus of the dataset on the direct tumor environment and its intersection with other tissue types. An example of this is displayed in Supplementary Figure 3 in which necrosis is correctly identified on the intersection with tumor tissue but not in the central necrotic patch.

No association was found between treatment response and the relative proportion of necrosis or blood vessels relative to viable tumor area (Supplementary Figure 4).

#### 3.2.3. Nuclei detection

The predictive value of detected nuclei was assessed only within manually annotated viable tumor areas, as tissue model segmentation outside these regions often resulted in substantial misclassification (see Supplementary Figure 3). These annotations have been checked by a pathologist and have previously been validated (Mark Schuiveling et al., 2025b). This approach allowed for annotations of TILs within both the intra-tumoral stroma and the tumor area.

Distribution analyses showed that lymphocytes were present in both tumor and stromal compartments, whereas tumor cells were largely confined to tumor regions. Stromal and intratumoral TIL densities were moderately correlated (Spearman ρ = 0.566 for track 1 and ρ = 0.589 for track 2, both p < 0.001). Models that incorporated tissue segmentation as input, including those by Torbati et al. and D. Adams, detected fewer tumor cells within stromal regions. In track 2, stromal cells and histiocytes were primarily localized to the tumor-associated stroma, while apoptotic cells were predominantly detected within tumor regions (Supplementary Figure 5).

The model as provided by A. Galdran identified a large number of melanophages, whereas the total number of nuclei detected was lower. This was confirmed by qualitative evaluation showing a substantial number of misclassifications, further evaluation demonstrated that the algorithms by Torbati et al and Lv et al. algorithms were able to detect plasma cells (Supplementary Figures 5 and 6).

##### 3.2.3.1 Nuclei detection and treatment outcomes

In univariable analyses, intra-tumoral lymphocytes showed the strongest association with a higher likelihood of treatment response across both track 1 and track 2, with odds ratios of around 1.34 per standard deviation (SD) increase across most algorithms. This association was weaker but remained significant for the algorithm by A. Galdran (OR 1.18, 95% CI 1.04–1.35). Lymphocytes in the intra-tumoral stroma were also associated with treatment response, with average odds ratios of approximately 1.24.

Plasma cells within the tumor were positively associated with treatment response in univariable analyses, whereas neutrophil proportions were not. Melanophages were negatively associated with treatment response in a subset of models.

In multivariable analyses including lymphocytes, plasma cell, and melanophages, only intra-tumoral lymphocyte proportion consistently remained associated with a higher likelihood of treatment response across participants. In the model by Torbati et al., stromal histiocytes also remained associated with response (Figure 2). No significant interaction effects between immune cell subsets were observed (data not shown).

**Figure 1.**
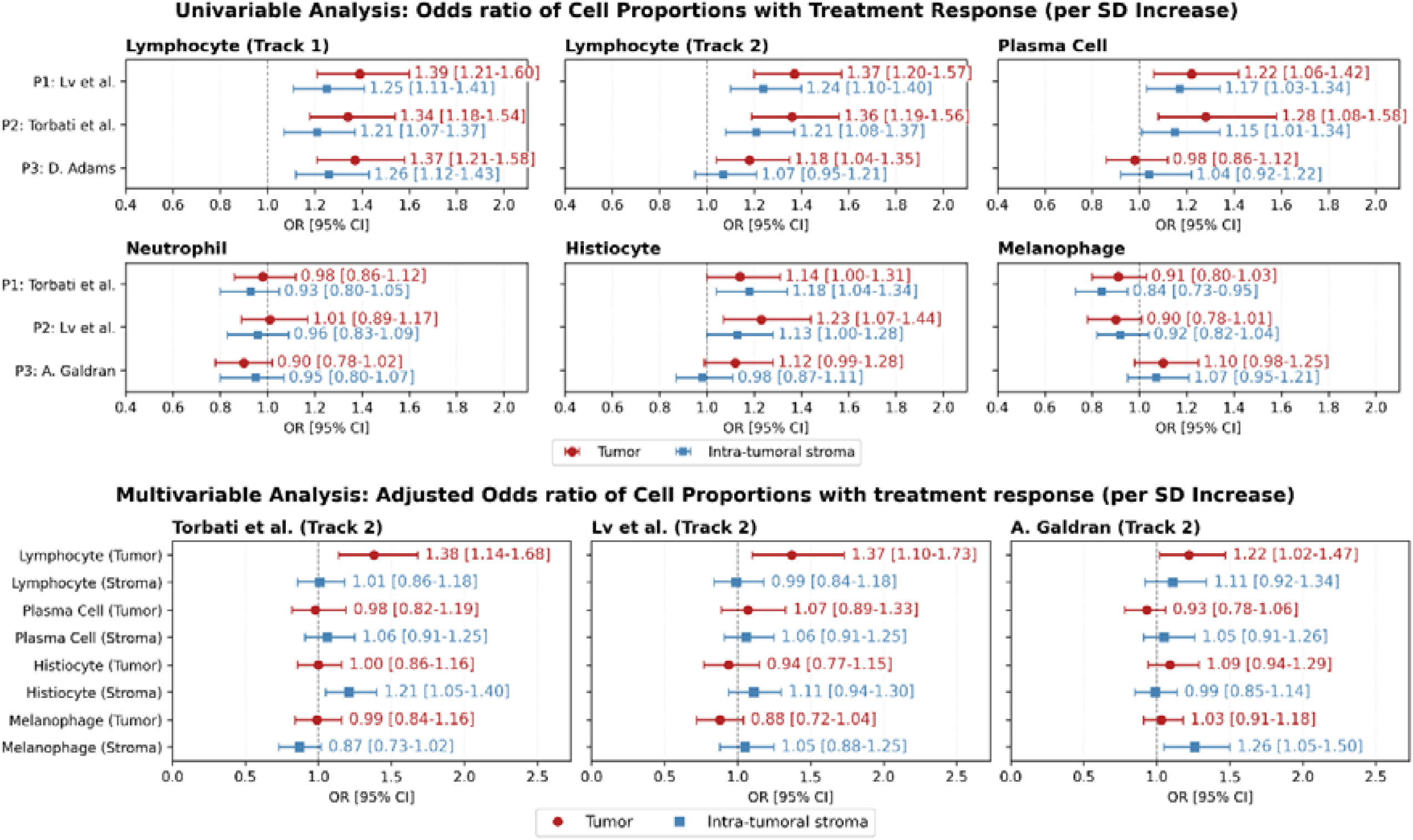
A. Univariable association of immune cell subsets increase per SD with treatment response B. Multivariable association of immune cell subsets increase per SD with treatment response.

**Figure 2.**
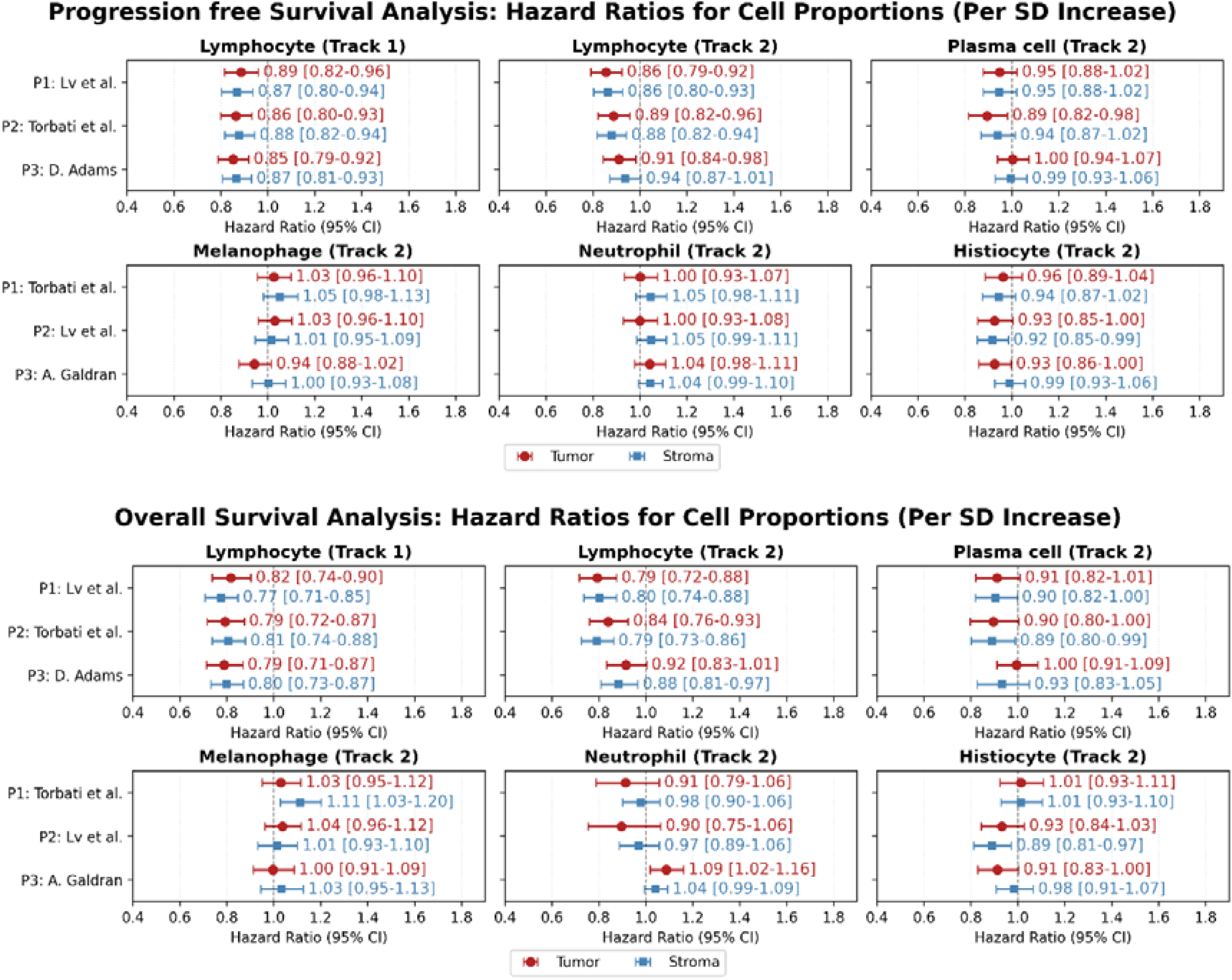
A. Association of immune cell subsets with progression free survival B. Association of immune cell subsets with overall survival

Lymphocytes were the only immune cell subset consistently associated with progression-free and overall survival, with similar effects observed in both tumor and stromal compartments (Figure 2). No consistent associations were observed between survival outcomes and the presence of plasma cells, melanophages, neutrophils, or histiocytes.

## 4. Discussion

This study evaluated the performance and clinical relevance of algorithms developed in the PUMA challenge. Two main findings were identified. First, the PUMA challenge led to clear improvements in tissue segmentation and nuclei detection. Second, intra-tumoral tumor-infiltrating lymphocytes as detected by the top submitted methods were the only consistent feature associated with response and survival to immune checkpoint inhibitors across participants and tracks.

The PUMA challenge demonstrated that it is possible to improve state of the art nuclei detection models resulting in improved immune cell characterization in melanoma histopathology. The highest average *F*_1_ score was acquired by Torbati et al. who used a multistage panoptic approach using both tissue masks and image data as input for nuclei detection. This resulted in improvement of location dependent nuclei classes such as apoptotic cells (which are mainly in necrotic areas) and epithelial cells (which are located only in the epithelium). Interestingly, Lv et al. and A. Galdran achieved similar average F1 scores using respectively class specific heads in a multi-headed architecture or improved balancing of training data. This resulted in higher detection performance of other, non-location dependent, nuclei classes such as melanophages, neutrophils or plasma cells.

Despite the improvement in nuclei detection, performance for other immune subsets detection remained modest. This likely reflects limitations of the reference dataset, as labels were based on visual classification on H&E, for which interobserver agreement is very low for several immune subtypes. A potential approach is to derive training labels by aligning H&E with immunohistochemical stains or molecular readouts, such as immunofluorescence or spatial transcriptomics. This approach is used in the STHELAR dataset (Giraud-Sauveur et al., 2025), yet even there, plasma-cell detection on H&E remains limited (F1 around 0.39), suggesting that some immune subsets may not be reliably separable from H&E-stained morphology alone.

In the second part of the manuscript we evaluated the correlation between spatially derived immune cell subsets and treatment outcomes. In univariable analyses, intra-tumoral and stromal plasma cells and histiocytes were associated with treatment response; however, these associations were no longer observed after adjustment for lymphocytes in multivariable models. Furthermore, no associations between plasma cells or histiocytes and survival outcomes were observed.

The absence of independent associations for plasma cells may partly reflect the limited sensitivity of morphology-based detection on H&E-stained tissue whereas our findings contrast with earlier research in non-small cell lung cancer in which plasma cells were associated with improved ICI outcomes (Patil et al., 2022). However, an important difference is that the study by Patil et al. measured plasma cell presence through RNA sequencing whereas this study only evaluated H&E-stained tissue resulting in limited detection of plasma cells. Alternatively, plasma cells and TILs may both capture the same broader inflamed microenvironment, necessary for an effective anti-tumor response (Blank et al., 2016). This could also explain why an univariable association was observed between histiocytes and response, an effect that was no longer present after adjustment for TILs.

Further exploratory analyses also demonstrated no correlation between necrosis or blood vessels and treatment outcomes. Previous studies have reported mixed results for these tissue features. In a pan-cancer cohort of patients treated with immune checkpoint inhibitors, Lee et al. reported that high endothelial cell (EC) presence was associated with poorer treatment outcomes (Lee et al., 2025). However, EC density varied strongly by tumor type, with the highest levels observed in renal cell carcinoma and hepatocellular carcinoma, and the lowest levels in pancreatic adenocarcinoma, melanoma, and cholangiocarcinoma. As pancreatic adenocarcinoma and cholangiocarcinoma are tumor types with generally low response rates to ICIs, this suggests that the reported association between EC presence and outcome may partly reflect differences between tumor types rather than a tumor-type–independent effect of ECs. Similarly, studies by Fa’ak et al. and our group in the same cohort reported that necrotic tumor phenotypes, with or without pigmentation, were associated with worse outcomes (Fa’ak et al., 2025; M. Schuiveling et al., 2025). This aligns with our findings, in which some models showed a negative association between melanophages and treatment response. However, we did not observe an association between necrosis itself and response. Qualitative assessment suggested that necrosis was not reliably identified by the tissue segmentation models, likely due to limited representation of necrotic tissue in the training data. Taken together, the lack of an observed association between necrosis and treatment response in this study should be interpreted with caution, as it does not provide sufficient evidence to exclude a potential relationship.

This study has several strengths. First, the PUMA dataset ranked among the top-performing datasets in a large-scale cross-dataset benchmark study (Torbati et al., 2026), supporting the robustness and quality of its annotations. Second, the PUMA dataset challenge algorithms were evaluated not only on a hidden test set but also on independent real-world whole-slide images, enabling assessment of generalizability beyond technical benchmarking. Third, the validation cohort was a large, multicenter cohort of patients with advanced melanoma treated with first-line ICIs. The cohort consisted out of prospectively collected clinical data and clinically relevant outcomes, which strengthens the translational relevance of the results.

This study also has several limitations. First, the PUMA training dataset contains some uncertainty in the ground truth annotations for nuclei and tissue classes, which is reflected by lower interobserver agreement and modest *F*_1_ scores for less common cell types. However, as stated earlier, even in datasets with more consistent ground truth based on spatial transcriptomics, detection performance for rare cell types remains low. This suggests that the observed classification performance of cells is likely close to what can be achieved using visual data from H&E-stained slides. Second, some patients were excluded from the clinical validation due to technical processing issues or the absence of a pre-treatment metastatic sample. Importantly, patient characteristics were similar between included and excluded patients, reducing the risk of selection bias. Finally, further validation and integration with clinical and molecular factors will be required before nuclei detection models can be applied in routine practice.

In conclusion, results from the PUMA challenge show that combining nuclei and tissue segmentation can improve cell detection and spatial analysis in melanoma histopathology. The findings indicate that immune cell location matters, with lymphocytes inside the tumor being more strongly associated with treatment response than those in the surrounding stroma. This spatial information allowed identification of clinically relevant associations, with intra-tumoral lymphocytes being the most consistent predictor of response to ICIs. In addition, the challenge led to clear improvements in tissue segmentation, especially for less common tissue classes, which may be useful for future applications in other settings.

## Data Availability

The training part of the PUMA dataset is publicly available. The preliminary test set and final test set are not publicly available as the PUMA challenge is a rolling challenge. The clinical dataset cannot be shared due to data sharing agreements with participating centers.

https://zenodo.org/records/15050523

## Acknowledgements

We would like to thank the Medical Imaging with Deep Learning (MIDL) foundation for their financial and technical support in organizing this challenge. We would also like to thank the Grand Challenge support team for their help in organizing this challenge. Furthermore, we thank Palga, the Dutch Nationwide Pathology Databank, for providing histopathological data and for their help in the collection of formalin-fixed paraffin-embedded tissue. Finally, we would like to thank all participants who joined the challenge. This research was funded by The Netherlands Organization for Health Research and Development (ZonMW, project number 848101007), by an unrestricted grant of Stichting Hanarth Fonds, The Netherlands, and by Philips.

## Contributors

All authors had responsibility for the decision to submit for publication.

M. Schuiveling and H. Liu contributed equally to this work.

M Veta, KPM Suijkerbuijk, and WAM Blokx contributed equally to this work.

**Concept and design**: Schuiveling, Liu, Veta, Suijkerbuijk, Blokx.

**Acquisition, analysis, or interpretation of data:** All authors.

**Drafting of the manuscript:** Schuiveling, Veta, Suijkerbuijk, Blokx, Liu

**Critical revision of the manuscript:** Schuiveling, van Duin, ter Maat, van der Weerd, van Eek, Liu, Hanusová, van den Berkmortel, Blank, Breimer, Burgers, Boers-Sonderen, van den Eertwegh, de Groot, Haanen, Hospers, Kapiteijn, Simkens, Westgeest, Schrader, Pluim, van Diest, Veta, Suijkerbuijk, Blokx. Lv, Zhu, Colin Tenorio, Chohan, Eastwood, Ahmed Raza, Torbati, Meshcheryakova, Mechtcheriakova, Mahbod, Adams, Galdran.

**Statistical analysis:** Schuiveling, Kapiteijn, Suijkerbuijk.

**Obtained funding:** Veta, Suijkerbuijk

**Administrative, technical, or material support:** Schuiveling, van Duin, ter Maat, van den Berkmortel, Burgers, van den Eertwegh, de Groot, Hospers, Kapiteijn, Piersma, Schrader, van Diest, van Eek, Liu, Hanusová

**Model development:** Lv, Zhu, Colin Tenorio, Chohan, Eastwood, Ahmed Raza, Torbati, Meshcheryakova, Mechtcheriakova, Mahbod, Adams, Galdran.

**Supervision:** Pluim, van Diest, Veta, Suijkerbuijk, Blokx.

## References

Bankhead, P., Loughrey, M.B., Fernández, J.A., Dombrowski, Y., McArt, D.G., Dunne, P.D., McQuaid, S., Gray, R.T., Murray, L.J., Coleman, H.G., James, J.A., Salto-Tellez, M., Hamilton, P.W., 2017. QuPath: Open source software for digital pathology image analysis. Sci Rep 7, 16878. 10.1038/s41598-017-17204-5

Baumann, E., Dislich, B., Rumberger, J.L., Nagtegaal, I.D., Martınez, M.R., Zlobec, I., 2024. HoVer-NeXt: A Fast Nuclei Segmentation and Classification Pipeline for Next Generation Histopathology. Medical Imaging with Deep Learning.

Blank, C.U., Haanen, J.B., Ribas, A., Schumacher, T.N., 2016. The “cancer immunogram.” Science 352, 658–660. 10.1126/science.aaf2834

Casparie, M., Tiebosch, A.T.M.G., Burger, G., Blauwgeers, H., van de Pol, A., van Krieken, J.H.J.M., Meijer, G.A., 2007. Pathology databanking and biobanking in The Netherlands, a central role for PALGA, the nationwide histopathology and cytopathology data network and archive. Cell Oncol 29, 19–24. 10.1155/2007/971816

Duin, I.A.J. van, Schuiveling, M., Maat, L.S. ter, Amsterdam, W.A.C. van, Berkmortel, F. van den, Boers-Sonderen, M., Groot, J.W.B. de, Hospers, G.A.P., Kapiteijn, E., Labots, M., Piersma, D., Schrader, A.M.R., Vreugdenhil, G., Westgeest, H., Veta, M., Blokx, W.A.M., Diest, P.J. van, Suijkerbuijk, K.P.M., 2024. Baseline tumor-infiltrating lymphocyte patterns and response to immune checkpoint inhibition in metastatic cutaneous melanoma. European Journal of Cancer 208. 10.1016/j.ejca.2024.114190

Eisenhauer, E.A., Therasse, P., Bogaerts, J., Schwartz, L.H., Sargent, D., Ford, R., Dancey, J., Arbuck, S., Gwyther, S., Mooney, M., Rubinstein, L., Shankar, L., Dodd, L., Kaplan, R., Lacombe, D., Verweij, J., 2009. New response evaluation criteria in solid tumours: Revised RECIST guideline (version 1.1). European Journal of Cancer, Response assessment in solid tumours (RECIST): Version 1.1 and supporting papers 45, 228–247. 10.1016/j.ejca.2008.10.026

Fa’ak, F., Coudray, N., Jour, G., Ibrahim, M., Illa-Bochaca, I., Qiu, S., Claudio Quiros, A., Yuan, K., Johnson, D.B., Rimm, D.L., Weber, J.S., Tsirigos, A., Osman, I., 2025. Artificial Intelligence Algorithm Predicts Response to Immune Checkpoint Inhibitors. Clin Cancer Res 31, 3526–3536. 10.1158/1078-0432.CCR-24-3720

Gershenwald, J.E., Scolyer, R.A., Hess, K.R., Sondak, V.K., Long, G.V., Ross, M.I., Lazar, A.J., Faries, M.B., Kirkwood, J.M., McArthur, G.A., Haydu, L.E., Eggermont, A.M.M., Flaherty, K.T., Balch, C.M., Thompson, J.F., for members of the American Joint Committee on Cancer Melanoma Expert Panel and the International Melanoma Database and Discovery Platform, 2017. Melanoma staging: Evidence-based changes in the American Joint Committee on Cancer eighth edition cancer staging manual. CA: A Cancer Journal for Clinicians 67, 472–492. 10.3322/caac.21409

Giraud-Sauveur, F., Blampey, Q., Benkirane, H., Marinello, A., Cournède, P.-H., Christodoulidis, S., 2025. STHELAR, a multi-tissue dataset linking spatial transcriptomics and histology for cell type annotation. 10.1101/2025.07.11.664123

Isensee, F., Jaeger, P.F., Kohl, S.A.A., Petersen, J., Maier-Hein, K.H., 2021. nnU-Net: a self-configuring method for deep learning-based biomedical image segmentation. Nat Methods 18, 203–211. 10.1038/s41592-020-01008-z

Jochems, A., Schouwenburg, M.G., Leeneman, B., Franken, M.G., van den Eertwegh, A.J.M., Haanen, J.B.A.G., Gelderblom, H., Uyl-de Groot, C.A., Aarts, M.J.B., van den Berkmortel, F.W.P.J., Blokx, W.A.M., Cardous-Ubbink, M.C., Groenewegen, G., de Groot, J.W.B., Hospers, G.A.P., Kapiteijn, E., Koornstra, R.H., Kruit, W.H., Louwman, M.W., Piersma, D., van Rijn, R.S., Ten Tije, A.J., Vreugdenhil, G., Wouters, M.W.J.M., van der Hoeven, J.J.M., 2017. Dutch Melanoma Treatment Registry: Quality assurance in the care of patients with metastatic melanoma in the Netherlands. Eur J Cancer 72, 156–165. 10.1016/j.ejca.2016.11.021

Kleczek, P., Jaworek-Korjakowska, J., Gorgon, M., 2020. A novel method for tissue segmentation in high-resolution H&E-stained histopathological whole-slide images. Computerized Medical Imaging and Graphics 79, 101686. 10.1016/j.compmedimag.2019.101686

Lee, S., Oh, J.W., Hwang, S., Shen, J., Park, S., Kim, H., Chae, Y.K., Lee, S.-H., Choi, Y.-L., Chung, J., Shin, J., Song, H., Valero Puche, A., Yoo, D., Lee, T., Oum, C., Kim, J., Ali, S.M., Ock, C.-Y., 2025. Deep learning–powered H&E whole-slide image analysis of endothelial cells to characterize tumor vascular environment and correlate treatment outcome to immunotherapy. J Clin Oncol 43, 2578–2578. 10.1200/JCO.2025.43.16_suppl.2578

Lv, J., Nasir, E.S., Xu, K., Jahanifar, M., Chohan, B.S., Elhaminia, B., Raza, S.E.A., 2025a. KongNet: A Multi-headed Deep Learning Model for Detection and Classification of Nuclei in Histopathology Images. 10.48550/arXiv.2510.23559

Lv, J., Zhu, Y., Tenorio, C.G.C., Chohan, B.S., Eastwood, M., Raza, S.E.A., 2025b. Leveraging Pathology Foundation Models for Panoptic Segmentation of Melanoma in H&E Images. 10.48550/arXiv.2507.13974

Maier-Hein, L., Reinke, A., Godau, P., Tizabi, M.D., Buettner, F., Christodoulou, E., Glocker, B., Isensee, F., Kleesiek, J., Kozubek, M., Reyes, M., Riegler, M.A., Wiesenfarth, M., Kavur, A.E., Sudre, C.H., Baumgartner, M., Eisenmann, M., Heckmann-Nötzel, D., Rädsch, T., Acion, L., Antonelli, M., Arbel, T., Bakas, S., Benis, A., Blaschko, M.B., Cardoso, M.J., Cheplygina, V., Cimini, B.A., Collins, G.S., Farahani, K., Ferrer, L., Galdran, A., van Ginneken, B., Haase, R., Hashimoto, D.A., Hoffman, M.M., Huisman, M., Jannin, P., Kahn, C.E., Kainmueller, D., Kainz, B., Karargyris, A., Karthikesalingam, A., Kofler, F., Kopp-Schneider, A., Kreshuk, A., Kurc, T., Landman, B.A., Litjens, G., Madani, A., Maier-Hein, K., Martel, A.L., Mattson, P., Meijering, E., Menze, B., Moons, K.G.M., Müller, H., Nichyporuk, B., Nickel, F., Petersen, J., Rajpoot, N., Rieke, N., Saez-Rodriguez, J., Sánchez, C.I., Shetty, S., van Smeden, M., Summers, R.M., Taha, A.A., Tiulpin, A., Tsaftaris, S.A., Van Calster, B., Varoquaux, G., Jäger, P.F., 2024. Metrics reloaded: recommendations for image analysis validation. Nat Methods 21, 195–212. 10.1038/s41592-023-02151-z

Patil, N.S., Nabet, B.Y., Müller, S., Koeppen, H., Zou, W., Giltnane, J., Au-Yeung, A., Srivats, S., Cheng, J.H., Takahashi, C., de Almeida, P.E., Chitre, A.S., Grogan, J.L., Rangell, L., Jayakar, S., Peterson, M., Hsia, A.W., O’Gorman, W.E., Ballinger, M., Banchereau, R., Shames, D.S., 2022. Intratumoral plasma cells predict outcomes to PD-L1 blockade in non-small cell lung cancer. Cancer Cell 40, 289–300.e4. 10.1016/j.ccell.2022.02.002

Rahib, L., Wehner, M.R., Matrisian, L.M., Nead, K.T., 2021. Estimated Projection of US Cancer Incidence and Death to 2040. JAMA Netw Open 4, e214708. 10.1001/jamanetworkopen.2021.4708

Schuiveling, M., 2024. Melanoma Histopathology Dataset with Tissue and Nuclei Annotations. 10.5281/zenodo.10940193

Schuiveling, M., Duin, I.V., Maat, L.S.T., Weerd, J.V. der, Berkmortel, F.V. den, Burgers, F., Boers-Sonderen, M., Labots, M., Groot, J.W.D., Haanen, J.B. a. G., Hospers, G., Kapiteijn, E., Piersma, D., Simkens, L.H., Westgeest, H.M., Diest, P.V., Pluim, J., Suijkerbuijk, K., Blokx, W., Veta, M., 2025. 36P Interpretable histomorphological subtypes linked to ICI response in advanced melanoma using AI-assisted histopathology analysis. ESMO Real World Data and Digital Oncology 10. 10.1016/j.esmorw.2025.100235

Schuiveling, Mark, Liu, H., Eek, D., Breimer, G.E., Suijkerbuijk, K.P.M., Blokx, W.A.M., Veta, M., 2025a. A novel dataset for nuclei and tissue segmentation in melanoma with baseline nuclei segmentation and tissue segmentation benchmarks. GigaScience 14, giaf011. 10.1093/gigascience/giaf011

Schuiveling, M., Liu, H., Eek, D., Breimer, G.E., Suijkerbuijk, K.P.M., Blokx, W.A.M., Veta, M., n.d. (2025) PUMA Challenge Baseline Track 1 (Version 1). [Computer software]. Software Heritage, https://archive.softwareheritage.org/swh:1:snp:c031cf421ddcfb62ee9a81227c7930ed132f250c;origin=https://github.com/tueimage/PUMA-challenge-eval-track1.

Schuiveling, M., Liu, H., Eek, D., Breimer, G.E., Suijkerbuijk, K.P.M., Blokx, W.A.M., Veta, M., n.d. (2025) PUMA Challenge Baseline Track 2 (Version 1). [Computer software]. Software Heritage, https://archive.softwareheritage.org/swh:1:snp:c031cf421ddcfb62ee9a81227c7930ed132f250c;origin=https://github.com/tueimage/PUMA-challenge-eval-track1.

Schuiveling, M., Liu, H., Eek, D., Breimer, G.E., Suijkerbuijk, K.P.M., Blokx, W.A.M., Veta, M., n.d. (2025) PUMA Challenge Evaluation Track 1 (Version 1). [Computer software]. Software Heritage, https://archive.softwareheritage.org/swh:1:snp:c031cf421ddcfb62ee9a81227c7930ed132f250c;origin=https://github.com/tueimage/PUMA-challenge-eval-track1.

Schuiveling, M., Liu, H., Eek, D., Breimer, G.E., Suijkerbuijk, K.P.M., Blokx, W.A.M., Veta, M., n.d. (2025) PUMA Challenge Evaluation Track 2 (Version 1). [Computer software]. Software Heritage, https://archive.softwareheritage.org/swh:1:snp:c031cf421ddcfb62ee9a81227c7930ed132f250c;origin=https://github.com/tueimage/PUMA-challenge-eval-track1.

Schuiveling, Mark, van Duin, I.A.J., Ter Maat, L.S., van der Weerd, J.C., Verheijden, R.J., van den Berkmortel, F., Blank, C.U., Breimer, G.E., Burgers, F.H., Boers-Sonderen, M., van den Eertwegh, A.J.M., de Groot, J.W.B., Haanen, J.B.A.G., Hospers, G.A.P., Kapiteijn, E., Piersma, D., Vreugdenhil, G., Westgeest, H., Schrader, A.M.R., Pluim, J.P.W., van Diest, P.J., Veta, M., Suijkerbuijk, K.P.M., Blokx, W.A.M., 2025b. Artificial Intelligence-Detected Tumor-Infiltrating Lymphocytes and Outcomes in Anti-PD-1-Based Treated Melanoma. JAMA Oncol e254072. 10.1001/jamaoncol.2025.4072

Shen, J., Choi, Y.-L., Lee, T., Kim, H., Chae, Y.K., Dulken, B.W., Bogdan, S., Huang, M., Fisher, G.A., Park, S., Lee, S.-H., Hwang, J.-E., Chung, J.-H., Kim, L., Song, H., Pereira, S., Shin, S., Lim, Y., Ahn, C.H., Kim, Seulki, Oum, C., Kim, Sukjun, Park, G., Song, S., Jung, W., Kim, Seokhwi, Bang, Y.-J., Mok, T.S.K., Ali, S.M., Ock, C.-Y., 2024. Inflamed immune phenotype predicts favorable clinical outcomes of immune checkpoint inhibitor therapy across multiple cancer types. J Immunother Cancer 12, e008339. 10.1136/jitc-2023-008339

Suijkerbuijk, K.P.M., van Eijs, M.J.M., van Wijk, F., Eggermont, A.M.M., 2024. Clinical and translational attributes of immune-related adverse events. Nature Cancer 5, 557–571. 10.1038/s43018-024-00730-3

Torbati, N., Meshcheryakova, A., Woitek, R., Hatamikia, S., Mechtcheriakova, D., Mahbod, A., 2026. NucFuseRank: Dataset Fusion and Performance Ranking for Nuclei Instance Segmentation. 10.48550/arXiv.2601.20104

Torbati, N., Meshcheryakova, A., Woitek, R., Hatamikia, S., Mechtcheriakova, D., Mahbod, A., 2025. A Multi-Stage Auto-Context Deep Learning Framework for Tissue and Nuclei Segmentation and Classification in H&E-Stained Histological Images of Advanced Melanoma. 10.48550/arXiv.2503.23958

Wolchok, J.D., Chiarion-Sileni, V., Gonzalez, R., Grob, J.-J., Rutkowski, P., Lao, C.D., Cowey, C.L., Schadendorf, D., Wagstaff, J., Dummer, R., Ferrucci, P.F., Smylie, M., Butler, M.O., Hill, A., Márquez-Rodas, I., Haanen, J.B.A.G., Guidoboni, M., Maio, M., Schöffski, P., Carlino, M.S., Lebbé, C., McArthur, G., Ascierto, P.A., Daniels, G.A., Long, G.V., Bas, T., Ritchings, C., Larkin, J., Hodi, F.S., 2022. Long-Term Outcomes With Nivolumab Plus Ipilimumab or Nivolumab Alone Versus Ipilimumab in Patients With Advanced Melanoma. J Clin Oncol 40, 127–137. 10.1200/JCO.21.02229

Zhang, S., Sun, L., Zuo, J., Feng, D., 2024. Tumor associated neutrophils governs tumor progression through an IL-10/STAT3/PD-L1 feedback signaling loop in lung cancer. Translational Oncology 40, 101866. 10.1016/j.tranon.2023.101866

