## Supplements for "Associations between spatial distribution of immune cell subsets and clinical outcomes in patients with advanced melanoma treated with immune checkpoint inhibitors: results from the PUMA challenge"

***Supplementary Table 1*** Nuclei segmentation performance of all teams including the baseline model in track 1 of the PUMA Grand Challenge. Results are sorted based on the final rank which is the average rank of the tissue and nuclei segmentation task.

| Final Rank | Rank | Team | Tumor | Lymphocyte | Other | Average *F*_1_ score |
| --- | --- | --- | --- | --- | --- | --- |
| 1 | 2 | Lv et al. | 0.84 [0.81–0.86] | 0.81 [0.79–0.83] | 0.58 [0.53–0.63] | 0.7439 [0.72 – 0.77] |
| 2 | 3 | Torbati et al. | 0.83 [0.81–0.85] | 0.79 [0.77–0.81] | 0.61 [0.56–0.66] | 0.7443 [0.72 – 0.77] |
| 3 | 1 | D. Adams | 0.85 [0.83–0.87] | 0.82 [0.79–0.84] | 0.60 [0.54–0.65] | 0.76 [0.73–0.78] |
| 4 | 4 | Biototem* | 0.81 [0.78–0.84] | 0.80 [0.77–0.82] | 0.54 [0.49–0.59] | 0.72 [0.69–0.74] |
| 5 | 5 | A. Galdran | 0.80 [0.78–0.82] | 0.78 [0.74–0.80] | 0.56 [0.52–0.60] | 0.71 [0.69–0.73] |
| 6 | 10 | Vison307 | 0.77 [0.75–0.79] | 0.77 [0.73–0.79] | 0.47 [0.44–0.51] | 0.67 [0.65–0.69] |
| 7 | 8 | gdeotale123 | 0.77 [0.75–0.79] | 0.74 [0.72–0.77] | 0.55 [0.51–0.60] | 0.69 [0.67–0.71] |
| 8 | 7 | *baseline* | 0.78 [0.75–0.80] | 0.79 [0.76–0.81] | 0.51 [0.46–0.56] | 0.69 [0.67–0.72] |
| 9 | 6 | hoerst | 0.81 [0.78–0.84] | 0.78 [0.75–0.81] | 0.50 [0.44–0.55] | 0.70 [0.67–0.72] |
| 10 | 9 | mlafarge* | 0.79 [0.76–0.81] | 0.78 [0.76–0.80] | 0.49 [0.44–0.53] | 0.69 [0.66–0.71] |
| 11 | 11 | aia_wt* | 0.79 [0.75–0.83] | 0.72 [0.65–0.77] | 0.51 [0.46–0.56] | 0.67 [0.63–0.71] |

* These participants did not add methodology description

***Supplementary Table 2*** Tissue segmentation performance of all teams including the baseline model in track 1 of the PUMA Grand Challenge. Results are sorted based on the final rank which is the average rank of the tissue and nuclei segmentation task.

| Final Rank | Rank | Team | Tumor | Stroma | Necrosis | Epidermis | Blood vessel | Average micro dice |
| --- | --- | --- | --- | --- | --- | --- | --- | --- |
| 1 | 1 | Lv et al. | 0.94 [0.92 - 0.95] | 0.84 [0.79 - 0.88] | 0.82 [0.02 - 0.93] | 0.86 [0.78 - 0.92] | 0.46 [0.35 - 0.54] | 0.78 [0.57 - 0.84] |
| 2 | 3 | Torbati et al. | 0.92 [0.89 - 0.95] | 0.81 [0.74 - 0.86] | 0.47 [0.00 - 0.70] | 0.87 [0.76 - 0.94] | 0.54 [0.46 - 0.61] | 0.72 [0.57 - 0.81] |
| 3 | 4 | D. Adams | 0.92 [0.89 - 0.94] | 0.82 [0.75 - 0.87] | 0.15 [0.00 - 0.56] | 0.81 [0.64 - 0.91] | 0.47 [0.38 - 0.55] | 0.63 [0.53 - 0.77] |
| 4 | 2 | Biototem* | 0.91 [0.89 - 0.94] | 0.81 [0.75 - 0.86] | 0.57 [0.00 - 0.90] | 0.86 [0.77 - 0.91] | 0.48 [0.39 - 0.55] | 0.73 [0.56 - 0.83] |
| 5 | 5 | A. Galdran | 0.92 [0.89 - 0.94] | 0.82 [0.77 - 0.86] | 0.16 [0.00 - 0.48] | 0.80 [0.64 - 0.89] | 0.41 [0.31 - 0.49] | 0.62 [0.52 - 0.73] |
| 6 | 6 | Vison307 | 0.92 [0.88 - 0.94] | 0.82 [0.76 - 0.87] | 0.10 [0.00 - 0.33] | 0.76 [0.58 - 0.89] | 0.52 [0.43 - 0.61] | 0.62 [0.53 - 0.73] |
| 7 | 9 | gdeotale123 | 0.91 [0.88 - 0.94] | 0.79 [0.71 - 0.84] | 0.02 [0.00 - 0.04] | 0.71 [0.46 - 0.86] | 0.35 [0.23 - 0.47] | 0.56 [0.46 - 0.63] |
| 8 | 10 | *baseline* | 0.91 [0.88 - 0.94] | 0.79 [0.71 - 0.84] | 0.02 [0.00 - 0.04] | 0.71 [0.46 - 0.86] | 0.35 [0.23 - 0.47] | 0.56 [0.46 - 0.63] |
| 9 | 7 | hoerst | 0.91 [0.88 - 0.93] | 0.80 [0.75 - 0.85] | 0.23 [0.00 - 0.43] | 0.75 [0.57 - 0.86] | 0.37 [0.26 - 0.47] | 0.61 [0.49 - 0.71] |
| 10 | 11 | mlafarge* | 0.90 [0.87 - 0.92] | 0.76 [0.71 - 0.80] | 0.07 [0.00 - 0.32] | 0.79 [0.64 - 0.87] | 0.44 [0.36 - 0.51] | 0.59 [0.51 - 0.68] |
| 11 | 8 | aia_wt* | 0.87 [0.83 - 0.90] | 0.62 [0.52 - 0.70] | 0.01 [0.00 - 0.07] | 0.69 [0.45 - 0.84] | 0.10 [0.04 - 0.17] | 0.46 [0.37 - 0.54] |

* These participants did not add methodology description

***Supplementary Table 3*** Nuclei segmentation performance of all teams including the baseline model in track 2 of the PUMA Grand Challenge. Results are sorted based on the final rank which is the average rank of the tissue and nuclei segmentation task.

| Final Rank | Rank | Team | Tumor | Lymphocyte | Plasma cell | Histiocyte | Neutrophil |
| --- | --- | --- | --- | --- | --- | --- | --- |
| 1 | 1 | Torbati et al. | 0.82 [0.80–0.84] | 0.75 [0.73–0.77] | 0.21 [0.10–0.31] | 0.46 [0.38–0.52] | 0.29 [0.15–0.46] |
| 2 | 3 | Lv et al. | 0.84 [0.81–0.86] | 0.81 [0.79–0.82] | 0.25 [0.07–0.41] | 0.43 [0.35–0.50] | 0.27 [0.10–0.53] |
| 3 | 2 | A. Galdran | 0.81 [0.79–0.83] | 0.80 [0.77–0.82] | 0.21 [0.02–0.39] | 0.47 [0.40–0.53] | 0.41 [0.22–0.62] |
| 4 | 7 | Vison307 | 0.74 [0.71–0.77] | 0.72 [0.68–0.75] | 0.13 [0.04–0.21] | 0.41 [0.35–0.45] | 0.20 [0.05–0.50] |
| 5 | 6 | Biototem* | 0.79 [0.75–0.82] | 0.77 [0.73–0.81] | 0.17 [0.05–0.27] | 0.31 [0.24–0.37] | 0.22 [0.15–0.36] |
| 6 | 8 | D. Adams | 0.85 [0.82–0.87] | 0.79 [0.76–0.82] | 0.00 [0.00–0.00] | 0.47 [0.40–0.53] | 0.06 [0.00–0.30] |
| 7 | 5 | mlafarge* | 0.79 [0.76–0.81] | 0.78 [0.76–0.80] | 0.31 [0.16–0.45] | 0.38 [0.32–0.44] | 0.26 [0.11–0.59] |
| 8 | 4 | gdeotale123 | 0.77 [0.74–0.79] | 0.75 [0.72–0.77] | 0.35 [0.21–0.43] | 0.42 [0.36–0.47] | 0.20 [0.08–0.39] |
| 9 | 9 | hoerst | 0.81 [0.78–0.83] | 0.77 [0.74–0.80] | 0.12 [0.03–0.21] | 0.39 [0.33–0.45] | 0.16 [0.05–0.31] |
| 10 | 11 | yangjian | 0.79 [0.76–0.81] | 0.74 [0.71–0.77] | 0.16 [0.07–0.24] | 0.35 [0.29–0.41] | 0.13 [0.02–0.38] |
| 11 | 12 | *baseline* | 0.73 [0.70–0.76] | 0.70 [0.66–0.73] | 0.10 [0.04–0.17] | 0.36 [0.30–0.41] | 0.17 [0.07–0.42] |
| 12 | 10 | aia_wt* | 0.79 [0.75–0.83] | 0.76 [0.70–0.81] | 0.24 [0.05–0.42] | 0.00 [0.00–0.00] | 0.36 [0.28–0.52] |
| Final Rank | **Rank** | **Team** | **Melanophage** | **Apoptosis** | **Endothelium** | **Epithelium** | **Stroma** |
| 1 | 1 | Torbati et al. | 0.38 [0.28–0.49] | 0.38 [0.24–0.51] | 0.44 [0.38–0.49] | 0.75 [0.62–0.81] | 0.41 [0.36–0.44] |
| 2 | 3 | Lv et al. | 0.44 [0.34–0.55] | 0.19 [0.12–0.28] | 0.44 [0.37–0.50] | 0.61 [0.45–0.69] | 0.38 [0.35–0.42] |
| 3 | 2 | A. Galdran | 0.45 [0.34–0.54] | 0.23 [0.18–0.29] | 0.36 [0.28–0.44] | 0.64 [0.49–0.72] | 0.40 [0.36–0.43] |
| 4 | 7 | Vison307 | 0.44 [0.34–0.53] | 0.22 [0.13–0.30] | 0.40 [0.33–0.46] | 0.60 [0.45–0.70] | 0.35 [0.31–0.38] |
| 5 | 6 | Biototem* | 0.33 [0.23–0.43] | 0.22 [0.14–0.30] | 0.43 [0.36–0.49] | 0.64 [0.45–0.73] | 0.34 [0.30–0.37] |
| 6 | 8 | D. Adams | 0.32 [0.22–0.43] | 0.08 [0.04–0.12] | 0.48 [0.41–0.55] | 0.72 [0.54–0.81] | 0.35 [0.30–0.38] |
| 7 | 5 | mlafarge* | 0.36 [0.28–0.45] | 0.16 [0.11–0.20] | 0.35 [0.29–0.41] | 0.64 [0.48–0.71] | 0.36 [0.33–0.40] |
| 8 | 4 | gdeotale123 | 0.40 [0.30–0.49] | 0.25 [0.17–0.34] | 0.37 [0.28–0.45] | 0.60 [0.38–0.72] | 0.40 [0.36–0.43] |
| 9 | 9 | hoerst | 0.35 [0.27–0.42] | 0.21 [0.14–0.28] | 0.31 [0.23–0.39] | 0.50 [0.28–0.63] | 0.34 [0.30–0.37] |
| 10 | 11 | yangjian | 0.32 [0.24–0.40] | 0.16 [0.12–0.21] | 0.34 [0.27–0.39] | 0.46 [0.27–0.57] | 0.35 [0.31–0.38] |
| 11 | 12 | *baseline* | 0.30 [0.21–0.37] | 0.06 [0.03–0.10] | 0.24 [0.19–0.29] | 0.03 [0.00–0.08] | 0.29 [0.26–0.32] |
| 12 | 10 | aia_wt* | 0.42 [0.31–0.52] | 0.23 [0.17–0.30] | 0.16 [0.11–0.22] | 0.55 [0.31–0.72] | 0.38 [0.34–0.41] |
| Final Rank | **Rank** | **Team** | **Average *F*_1_ score** |  |  |  |  |
| 1 | 1 | Torbati et al. | 0.49 [0.46–0.51] |  |  |  |  |
| 2 | 3 | Lv et al. | 0.47 [0.43–0.51] |  |  |  |  |
| 3 | 2 | A. Galdran | 0.48 [0.44–0.51] |  |  |  |  |
| 4 | 7 | Vison307 | 0.42 [0.39–0.46] |  |  |  |  |
| 5 | 6 | Biototem* | 0.42 [0.39–0.45] |  |  |  |  |
| 6 | 8 | D. Adams | 0.41 [0.38–0.44] |  |  |  |  |
| 7 | 5 | mlafarge* | 0.44 [0.41–0.48] |  |  |  |  |
| 8 | 4 | gdeotale123 | 0.45 [0.41–0.48] |  |  |  |  |
| 9 | 9 | hoerst | 0.39 [0.36–0.43] |  |  |  |  |
| 10 | 11 | yangjian | 0.38 [0.35–0.41] |  |  |  |  |
| 11 | 12 | *baseline* | 0.30 [0.28–0.33] |  |  |  |  |
| 12 | 10 | aia_wt* | 0.39 [0.35–0.43] |  |  |  |  |

* These participants did not add methodology description

***Supplementary Table 4*** Tissue segmentation performance of all teams including the baseline model in track 2 of the PUMA Grand Challenge. Results are sorted based on the final rank which is the average rank of the tissue and nuclei segmentation task.

| Final Rank | Rank | Team | Tumor | Stroma | Necrosis | Epidermis | Blood vessel | Average micro dice |
| --- | --- | --- | --- | --- | --- | --- | --- | --- |
| 1 | 1 | Torbati et al. | 0.92 [0.90 - 0.95] | 0.81 [0.74 - 0.86] | 0.75 [0.00 - 0.88] | 0.87 [0.76 - 0.94] | 0.54 [0.46 - 0.61] | 0.78 [0.57 - 0.85] |
| 2 | 2 | Lv et al. | 0.94 [0.92 - 0.95] | 0.84 [0.79 - 0.88] | 0.82 [0.02 - 0.93] | 0.86 [0.78 - 0.92] | 0.46 [0.35 - 0.54] | 0.78 [0.57 - 0.84] |
| 3 | 6 | A. Galdran | 0.92 [0.89 - 0.94] | 0.82 [0.77 - 0.86] | 0.16 [0.00 - 0.48] | 0.80 [0.64 - 0.89] | 0.41 [0.31 - 0.49] | 0.62 [0.52 - 0.73] |
| 4 | 3 | Vison307 | 0.92 [0.88 - 0.94] | 0.82 [0.76 - 0.87] | 0.10 [0.00 - 0.33] | 0.76 [0.58 - 0.89] | 0.52 [0.43 - 0.61] | 0.62 [0.53 - 0.73] |
| 5 | 4 | Biototem* | 0.91 [0.89 - 0.94] | 0.81 [0.75 - 0.86] | 0.57 [0.00 - 0.90] | 0.86 [0.77 - 0.91] | 0.48 [0.39 - 0.55] | 0.73 [0.56 - 0.83] |
| 6 | 5 | D. Adams | 0.92 [0.89 - 0.94] | 0.82 [0.75 - 0.87] | 0.15 [0.00 - 0.56] | 0.81 [0.64 - 0.91] | 0.47 [0.38 - 0.55] | 0.63 [0.53 - 0.77] |
| 7 | 9 | mlafarge* | 0.90 [0.87 - 0.92] | 0.76 [0.71 - 0.80] | 0.08 [0.00 - 0.32] | 0.79 [0.64 - 0.87] | 0.44 [0.36 - 0.51] | 0.59 [0.51 - 0.68] |
| 8 | 8 | gdeotale123 | 0.91 [0.88 - 0.94] | 0.79 [0.71 - 0.84] | 0.02 [0.00 - 0.04] | 0.71 [0.46 - 0.86] | 0.35 [0.23 - 0.47] | 0.56 [0.46 - 0.63] |
| 9 | 7 | hoerst | 0.91 [0.88 - 0.93] | 0.80 [0.75 - 0.85] | 0.23 [0.00 - 0.43] | 0.75 [0.57 - 0.86] | 0.37 [0.26 - 0.47] | 0.61 [0.49 - 0.71] |
| 10 | 11 | yangjian | 0.88 [0.83 - 0.91] | 0.78 [0.70 - 0.83] | 0.00 [0.00 - 0.01] | 0.71 [0.46 - 0.86] | 0.35 [0.23 - 0.47] | 0.55 [0.44 - 0.62] |
| 11 | 10 | baseline | 0.91 [0.88 - 0.94] | 0.79 [0.71 - 0.84] | 0.02 [0.00 - 0.04] | 0.71 [0.46 - 0.86] | 0.35 [0.23 - 0.47] | 0.56 [0.46 - 0.63] |
| 12 | 11 | aia_wt* | 0.87 [0.83 - 0.90] | 0.62 [0.52 - 0.70] | 0.01 [0.00 - 0.07] | 0.69 [0.45 - 0.84] | 0.10 [0.04 - 0.17] | 0.46 [0.37 - 0.54] |

* These participants did not add methodology description

***Supplementary Table 5*** *Baseline patient characteristics of included and excluded patients*

|  |  | Included N=1102 | Excluded N=747 |
| --- | --- | --- | --- |
| Age, median [Q1,Q3] |  | 67.0 [57.0,74.0] | 67.0 [56.0,75.5] |
| Sex, n (%) | Female | 401 (36.4) | 309 (41.4) |
|  | Male | 701 (63.6) | 438 (58.6) |
| Therapy type | Anti-PD1 | 704 (63.9) | 487 (65.2) |
|  | Anti-PD1 + anti-CTLA4 | 398 (36.1) | 260 (34.8) |
| WHO performance stage, n (%) | WHO 0 | 577 (52.4) | 411 (55.0) |
|  | WHO 1 | 382 (34.7) | 254 (34.0) |
|  | WHO 2 or above | 78 (7.1) | 55 (7.4) |
|  | Missing | 65 (5.9) | 27 (3.6) |
| Disease Stage  (AJCC 8th Edition), n (%) | IIIC | 97 (8.8) | 69 (9.2) |
|  | M1a | 95 (8.6) | 66 (8.8) |
|  | M1b | 164 (14.9) | 119 (15.9) |
|  | M1c | 485 (44.0) | 319 (42.7) |
|  | M1d with non-symptomatic brain metastasis | 169 (15.3) | 94 (12.6) |
|  | M1d with symptomatic brain metastasis | 92 (8.3) | 80 (10.7) |
| BRAF V600E/K Mutation, n (%) | Mutated | 359 (32.6) | 243 (32.5) |
|  | Wildtype | 630 (57.2) | 423 (56.6) |
|  | Missing | 113 (10.3) | 81 (10.8) |
| LDH levels, n (%) | Normal | 756 (68.6) | 511 (68.4) |
|  | 1-2 x ULN | 256 (23.2) | 167 (22.4) |
|  | >2 x ULN | 78 (7.1) | 60 (8.0) |
|  | Missing | 12 (1.1) | 9 (1.2) |
| Objective response | Responder | 631 (57.3) | 405 (54.2) |
|  | Non-responder | 471 (42.7) | 342 (45.8) |

ULN = Upper limit of normal

**Supplementary figures**

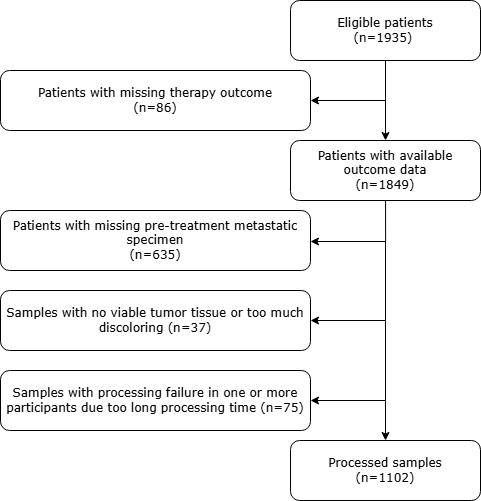

***Supplementary figure 1*** Flowchart of study population

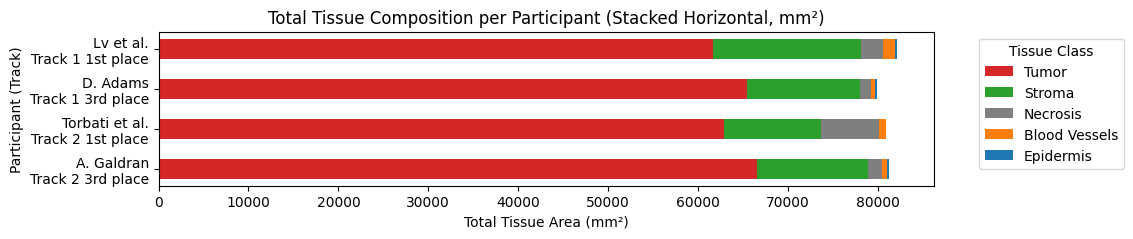

***Supplementary figure 2*** Distribution of tissue component areas in the validation dataset.

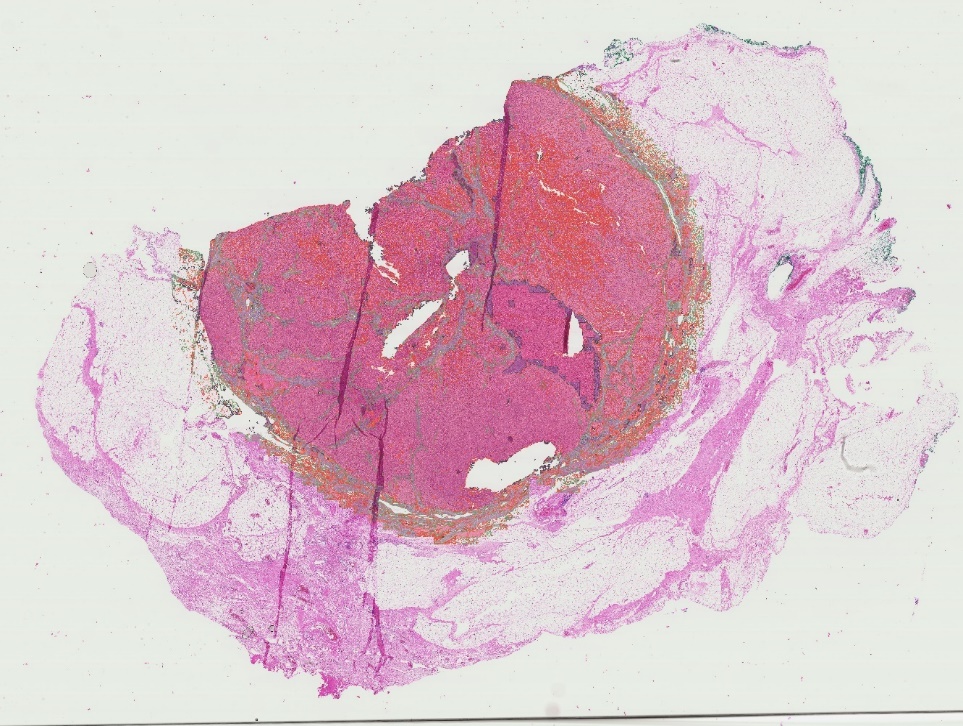

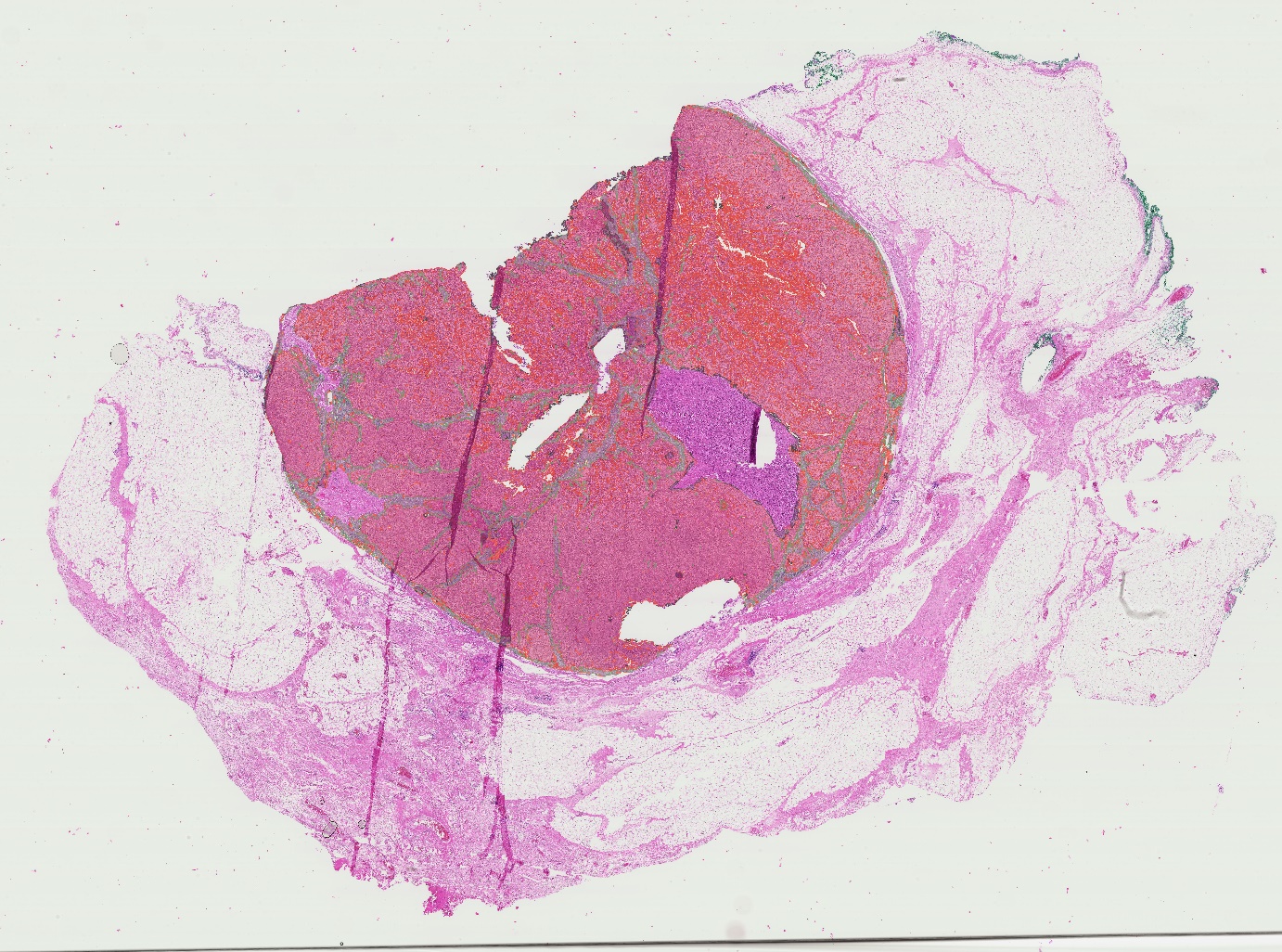

**B**

**A**

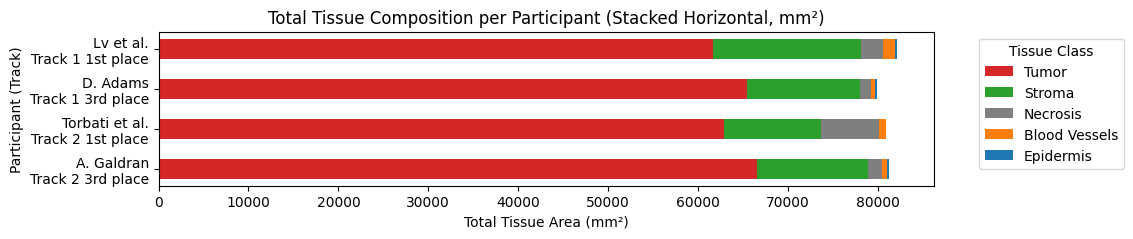

***Supplementary Figure 3*.** Melanoma metastases in a lymph node with tissue segmentation overlay.
Panel A shows the original segmentation overlay from the algorithm of Torbati et al. In this segmentation the large central necrotic part, as indicated with a white arrows, is mainly classified as tumor (in red).

Panel B demonstrates the segmentation result after filtering for manually annotated tumor area demonstrating only tumor and intra-tumoral stroma as included annotated areas.

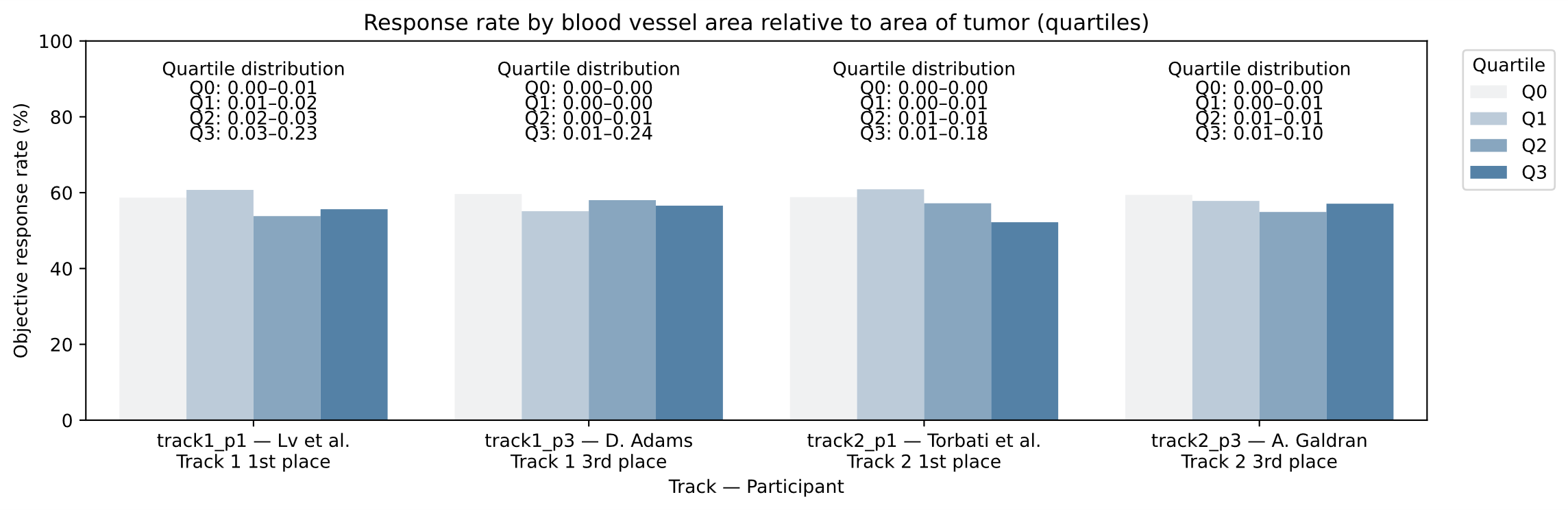
A

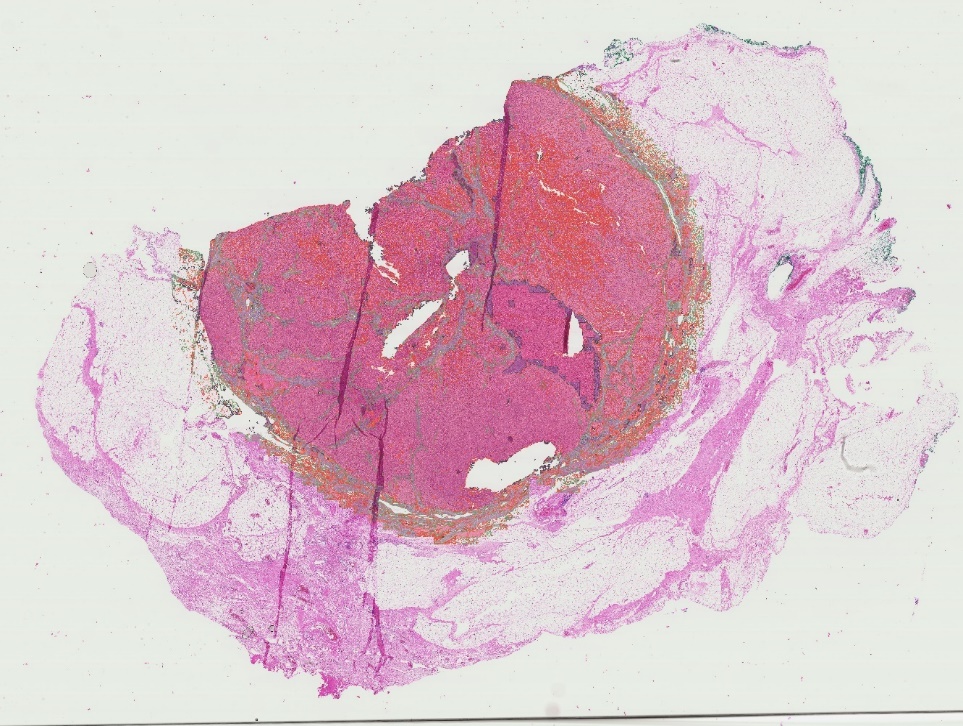

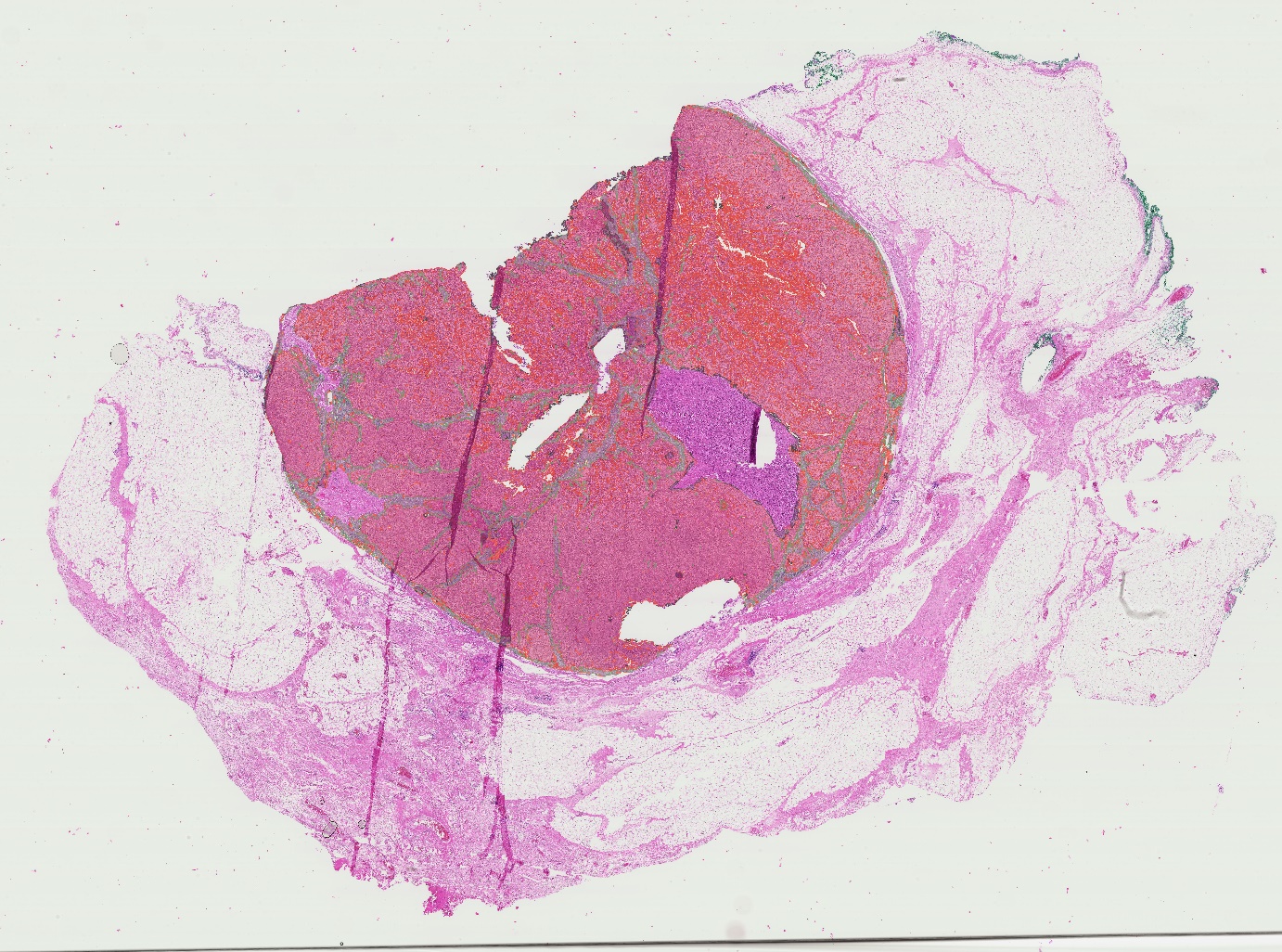

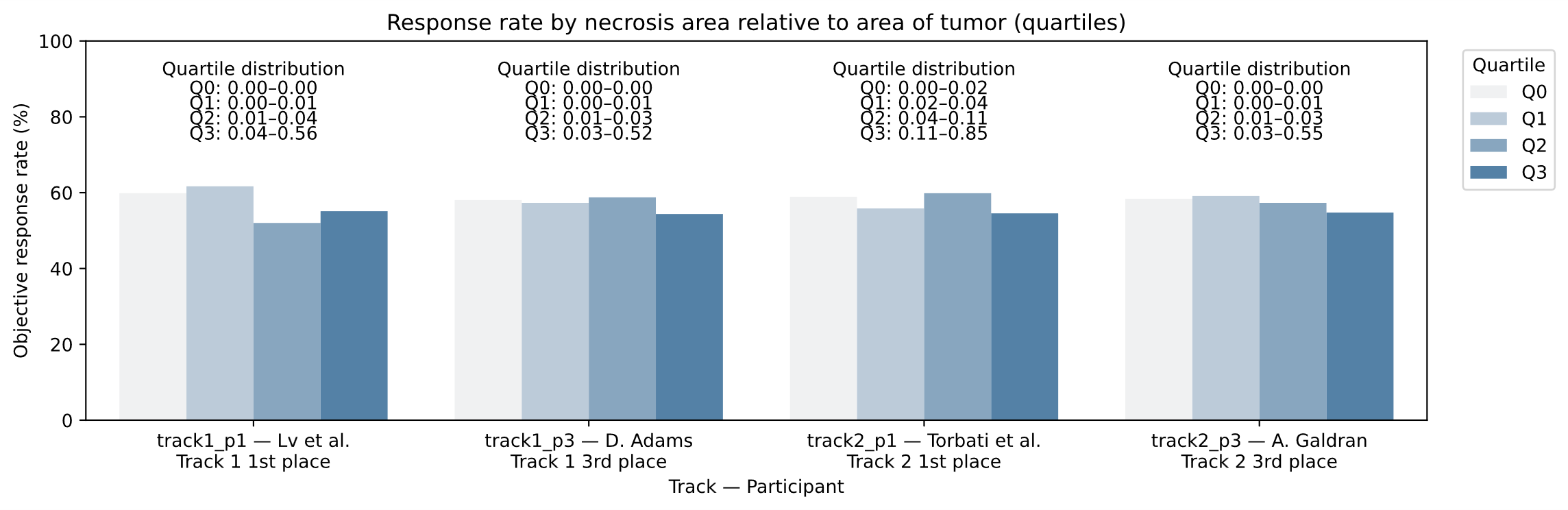
B

***Supplementary Figure 4*.** Objective response rate (ORR) across quartiles of blood vessel (panel A) and necrosis (panel B) proportion relative to tumor area for each participant–track combination.

Whole-slide images were stratified into quartiles based on the proportion of blood vessel area or necrotic tissue relative to total tumor area. Bars represent the mean ORR within each quartile, with color intensity reflecting increasing tissue proportions (Q0–Q3). Above each bar group, the distribution ranges for the corresponding quartile are shown. Across participants, no consistent trend was observed, and neither higher blood vessel proportions nor higher necrosis proportions showed a statistically significant association with treatment response.

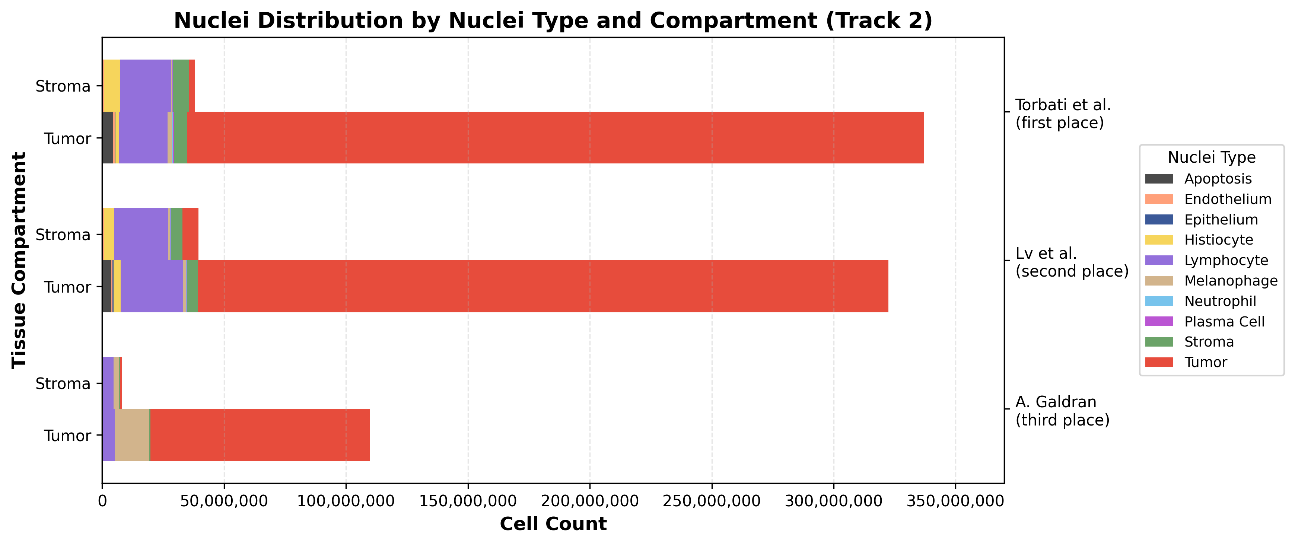

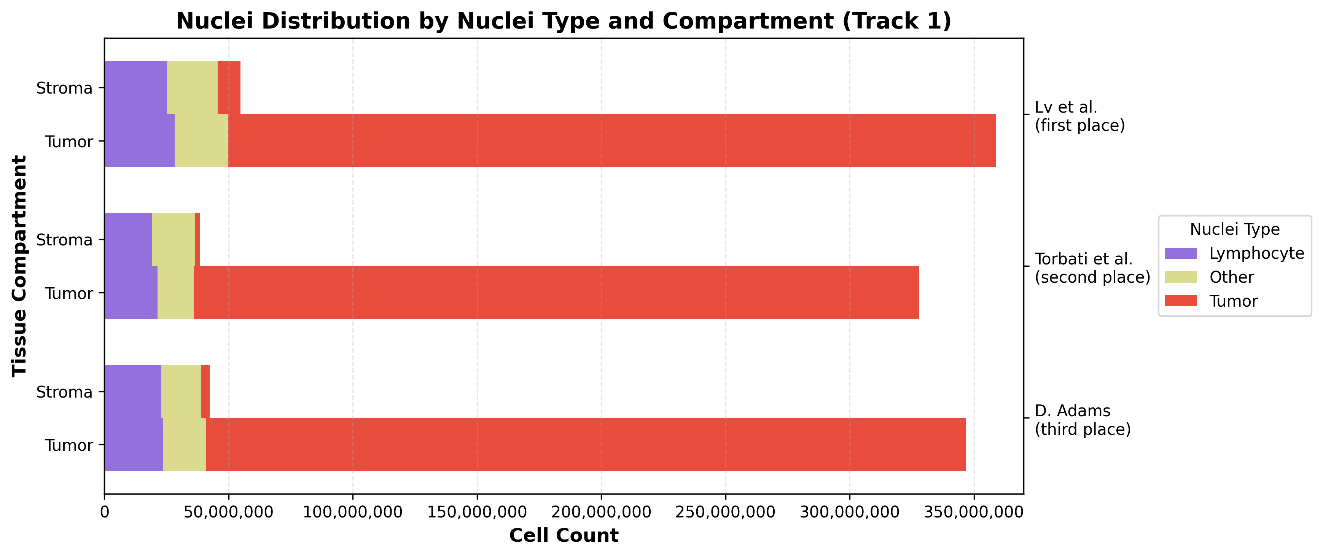
***Supplementary figure 5*** Distribution of nuclei detections in the validation dataset. Differences between the track 1 and track 2 for the algorithms from Lv et al. are due to differences in the processing pipelines. Track 2 was run using KongNet (Lv et al., 2025b), an optimized whole slide image nuclei segmentation model from Lv et al. which uses the same weights and setup as the PUMA challenge. However, post-processing of this model is based on a different non-maximum suppression threshold when compared to the PUMA challenge validation pipeline.

**A**

**B**

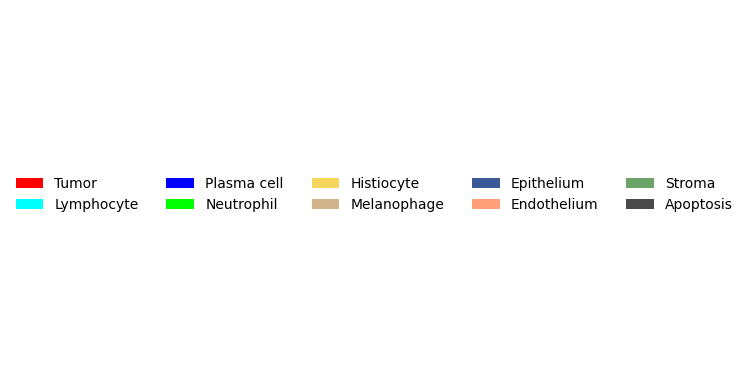

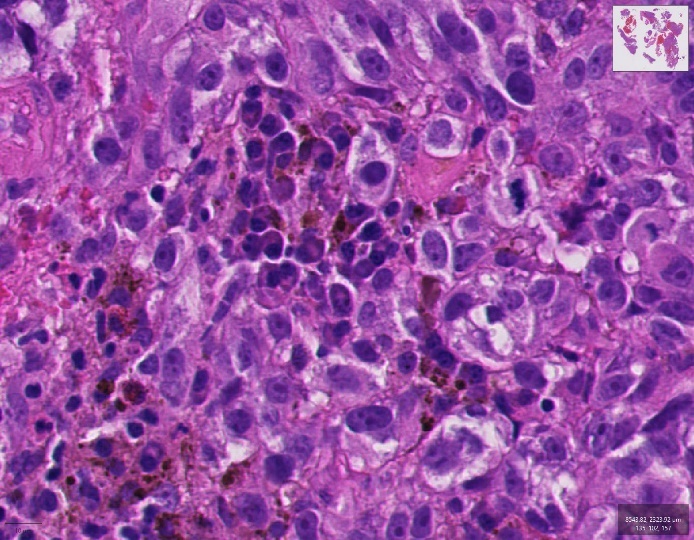

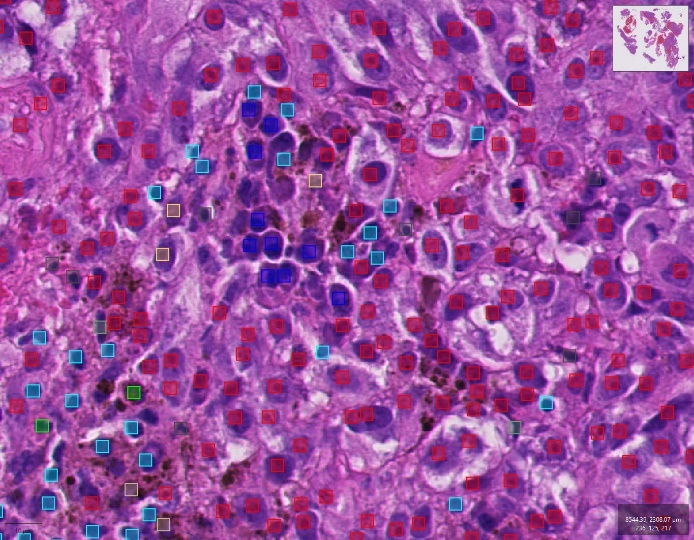

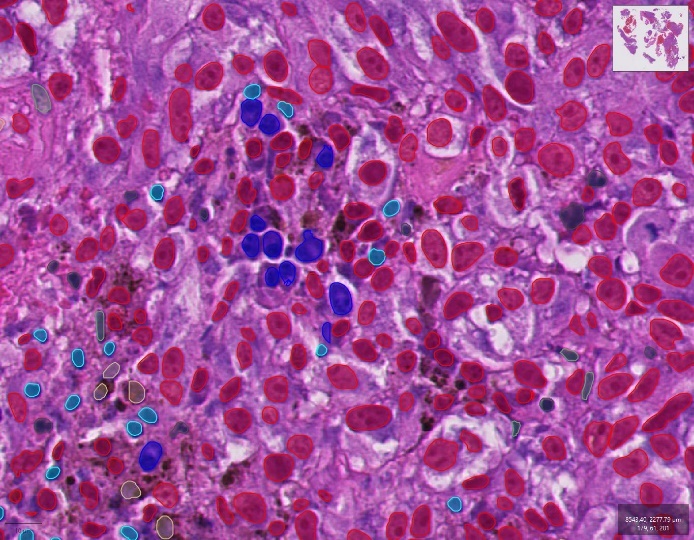

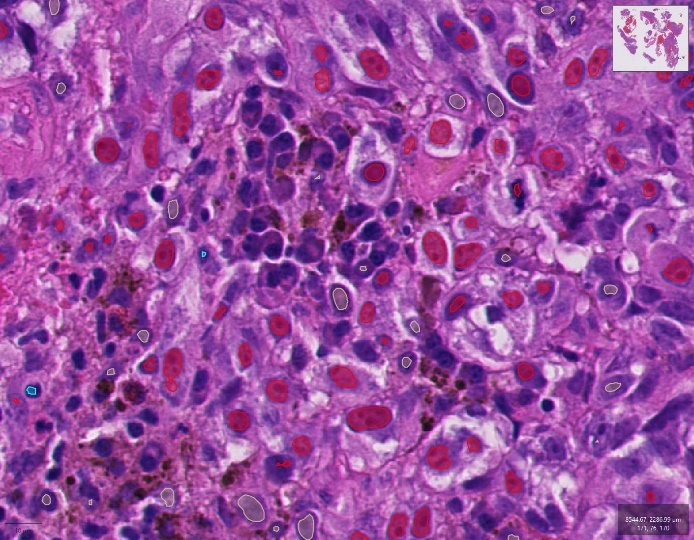

**D**

**C**

***Supplementary figure 6*** Example outputs from track 2 nuclei detection models. Panel A shows the original tissue region upon H&E-staining, B the results from Torbati et al., C from Lv et al., and D from A. Galdran*.*

**Supplementary methods**

***Appendix A.1. Method 1: TIAKong***

Team name: TIAKong

Authors: Jiaqi Lv, Yijie Zhu, Carmen Guadalupe Colin Tenorio, Brinder Singh Chohan, Mark Eastwood, Shan E Ahmed Raza

Affiliations: University of Warwick, University Hospitals of Derby and Burton NHS Foundation Trust, Medical University of Vienna, TissueGnostics GmbH

***Introduction***

Lv et al. used separate approaches for tissue and nuclei segmentation.
Tissue segmentation was done by leveraging Virchow2 (Paige, NYC, USA and Microsoft Research, Cambridge, MA USA, n.d.), a pathology foundation model trained on 3.1 million histopathology images as a feature extractor. These features are fused with the original RGB images and subsequently processed by an encoder-decoder segmentation network (Efficient-UNet) to produce accurate segmentation maps.(Lv et al., 2025c)

Nuclei detection was performed with KongNet; a multi-headed deep learning architecture in which a shared feature extractor combined with class-specific prediction heads, allowing separate optimization for different nuclei types and reducing interference between visually similar cell classes (Lv et al., 2025a).

***Pre-processing***

- Each image and its corresponding mask of shape 3×1024×1024 (*C*×*H*×*W*) pixels at 40× magnification are downsampled to 3×224×224 pixels using bilinear interpolation.
- Weighted sampling strategy is implemented. Patches containing more tissue of rare classes are assigned higher sampling probabilities.

***Data augmentation***

- Random RGB shift, random hue saturation and value (HSV) shift, Gaussian blur and sharpening, image compression, random brightness and contrast adjustments, random shifts and scaling, random 90-degree rotations and horizontal/vertical flips.

***Segmentation task method***

- Patch Token Extraction: The downsampled image X is passed to Virchow2, which produces a sequence of patch tokens T of dimension 1280×256.
- Permutation: T is rearranged into a multidimensional grid T’ of shape 1280×16×16. This facilitates convolution operations.
- Progressive Transposed Convolution (PTC) Module: T’ is progressively upsampled by the PTC Module. The output is a 5×224×224 spatially resolved feature map, T”.
- Feature Concatenation: T” is concatenated with the original image patch X, producing X’, a tensor of dimension 8×224×224. This step preserves both token-based features from Virchow2 and raw context from the input image.
- EfficientUNet Segmentation: X’ is passed to EfficientUNet, an encoderdecoder network, with an EfficientNetV2-M backbone. It outputs a segmentation map of dimension 5×224×224, corresponding to five target tissue classes.
- Dual Stage Loss: Loss is computed at two stages, after the PTC Module (LPTC) and at the final output stage (LOutput). LPTC and LOutput are combined using a weighted sum.

***Architecture***

- PTC module: this module consists of three convolution layers with sigmoid activations, which upsample patch tokens from 16×16 to 224×224 while reducing the channel dimension from 1280 to 5.
- Efficient-UNet: this is constructed with an encoder–decoder network with U-Net–style skip connections (Ronneberger et al., 2015) replacing the encoder with an EfficientNetV2-M (Tan and Le, 2021) backbone pre-trained on ImageNet (Russakovsky et al., 2015) , and added Spatial and Channel Squeeze & Excitation (SCSE) modules (Roy et al., 2018) into the decoder blocks.
- Dual Stage Loss: the loss function is a combination of Dice loss and Focal loss (DiceFL), with a larger weighting on Dice loss.

***Post-processing***

- To remove small holes and spurious objects, morphological opening followed by morphological closing is performed using a circular kernel of size 13 pixels.

***Results***

- Track1: Average F1 score: 0.74. Average micro dice score: 0.78. Final rank: 1.
- Track2: Average F1 score: 0.47. Average micro dice score: 0.78. Final rank: 2.

***Processing in validation study***

- Tissue annotations were created once as both tracks used the same model and weights
- Nuclei annotations for track 1 were created using the framework of the PUMA challenge validation.
- Nuclei annotations for track 2 were created using the released whole slide image (WSI) Inference pipeline for KongNet models (“Jiaqi-Lv/KongNet_Inference_Main: Main Github repository for KongNet Inference Code,” n.d.) resulting in a significant speedup of the pipeline.

***Appendix A.2. Method 2: LSM***

Team name: LSM

Authors: Nima Torbati, Anastasia Meshcheryakova, Diana Mechtcheriakova, and Amirreza Mahbod

Affiliations: Research Center for Medical Image Analysis and Artificial Intelligence, Department of Medicine, Faculty of Medicine and Dentistry, Danube Private University, 3500 Krems an der Donau, Austria  
Department of Pathophysiology and Allergy Research, Center of Pathophysiology, Infectiology and Immunology, Medical University of Vienna, Vienna, 1090, Austria
Comprehensive Center for AI in Medicine (CAIM), Medical University of Vienna, Vienna, 1090, Austria


***Introduction***

A novel multi-stage deep learning approach is proposed by combining tissue and nuclei information in a unified framework based on the autocontext concept to perform segmentation and classification in histological images of melanoma. (Torbati et al., 2025)

***Pre-processing***

- Models are Pre-trained on PanopTILs dataset (Liu et al., 2024).

***Data augmentation***

- Shape-based transformations (flip, rotation, scaling) and intensity-based adjustments (hue, saturation, brightness, contrast) are applied.

***Segmentation task method***

- Stage 1: a classifier is designed to determine the input image type, distinguishing between primary and metastatic frames.
- Stage 2: two SegFormer models are trained separately on the primary and metastatic subsets for initial tissue segmentation. For blood vessel prediction, the outputs from SegFormer are combined with the corresponding U-Net predictions. For epidermis and necrosis, the final result is obtained by averaging the predictions from both the SegFormer and U-Net models.
- Stage 3: the tissue segmentation from Stage 2 is incorporated as a fourth input channel, and a U-Net++ model is trained to predict the nuclei class map. The final instance class is determined using a simple majority-voting strategy. Nuclei instance masks are then generated using the pre-trained HoVer-NeXt model.
- Stage 4: tissue segmentation is refined using the nuclei masks produced in the stage 3.

***Architecture***

- Stage 1: SegFormer (Xie et al., 2021) (B2 variant) with ImageNet pre-trained weights (Russakovsky et al., 2015).
- Stage 2: SegFormer and U-Net (Ronneberger et al., 2015) with a pre-trained ResNet34 (He et al., 2016) encoder.
- Stage 3: UNet++ (Zhou et al., 2018) with a pretrained ResNet50 (He et al., 2016) encoder.
- Stage 4: Pre-trained HoVer-Next (Baumann et al., 2024) model, based on the PUMA training dataset(Schuiveling et al., 2024), is provided by the challenge organizers.

***Post-processing***

- Tissue: incorporate predictions from stage 1 when there is an overlap between the stage 1 and stage 4 predictions for necrotic tissue (only in Track 2).
- Nuclei: replace the instance segmentation results at image borders with the corresponding class map results.

***Results***

- Track1: Average F1 score: 0.74. Average micro dice score: 0.72. Final rank: 2.
- Track2: Average F1 score: 0.49. Average micro dice score: 0.78. Final rank: 1.

***Processing in validation study***

- Tissue annotations were created once due to very long processing time. The best performing model was chosen for the evaluation of both tracks (the tissue segmentation model from track 2).
- Nuclei segmentations were used from a previous assessment of the validation cohort with the Hover-NeXt model. The problem of incorrect annotations at the edge of a tile present at tile inference are not present on whole slide image inference due to the use of 512 pixel wide overlapping regions to check for half-instance segmentation masks. Classification was done through the majority voting approach as used in the multi-stage approach.

***Appendix A.3. Method 3: rictoo***

Team name: rictoo

Authors: Daniel Adams

Affiliation: The Cancer Epidemiology and Prevention Research Unit, The Institute of Cancer Research, London and Imperial College London, London, UK

Division of Genetics and Epidemiology, The Institute of Cancer Research, London, UK

***Introduction***

HoverLink, is proposed for joint tissue and nuclei segmentation. The model consists of two parallel branches. (Adams, 2026)

***Pre-processing***

- A 2048×2048 crop at the original 40× magnification is extracted from the larger ROI, containing the 1024×1024 annotated region at its center along with 512 pixels of surrounding real image content.
- Weighted sampling is applied during training, where sample weights are inversely proportional to the area of each tissue class.
- Images are normalized per RGB channel with ImageNet mean [0.485, 0.456, 0.406] and standard deviation [0.229, 0.224, 0.225].

***Data augmentation***

- Spatial augmentations, including mirroring, translation, scaling, zooming, rotation, shearing, and elastic deformations, color jitter are applied. These augmentations are performed on the full 2048×2048 crop before extracting the 1024×1024 center region for model input.

***Segmentation task method***

- Track 1 model (3-class nuclear output) trained for 250 epochs.
- Track 2 model (10-class nuclear output) trained for 250 epochs.

• Hybrid Track 1 model (250/350 epochs):

– The tissue branch is trained for 250 epochs, after which it is frozen.

– The nuclear branch continued training for an additional 100 epochs (totaling 350 epochs).

**Architecture**

- The first branch (A) performs tissue segmentation, while the second branch (B), based on the HoVerNeXt architecture, focuses on nuclear segmentation.
- Branches (A) and (B) are parallel U-Net [2] with ConvNeXt-Base backbones pretrained on ImageNet (Russakovsky et al., 2015)
- HoverLink uses cross-branch feature concatenation, where intermediate bottleneck features from A are concatenated with B.
- A learned feature transformation matrix refines the final nuclei segmentation by weighting per-pixel features from A and integrating them into the final activation of B.

***Post-processing***

- For tissue segmentation, the tissue outputs from all three models are aggregated in both Track 1 and Track 2.

• For nuclear segmentation:

– Track 1 used the 250/350 hybrid model for nuclear output (3-class nuclear segmentation).

– Track 2 used its own 250-epoch model for nuclear output (10-class nuclear segmentation).

***Results***

- Track1: Average F1 score: 0.76. Average micro dice score: 0.63. Final rank: 3.
- Track2: Average F1 score: 0.41. Average micro dice score: 0.63. Final rank: 6.

***Processing in validation cohort***

No specific changes were implemented for validation of the algorithm.

***Appendix A.4. Method 4: agaldran***

Team name: agaldran

Authors: Adrian Galdran

Affiliation: Tecnalia, Spain

***Introduction***

A standard semantic segmentation approach is used, consisting of multi-fold training of an encoder–decoder network.

***Pre-processing***

- Data splits: a subset of the entire dataset is selected that would have eight images per category, and for each category those images would have presence of it. The remaining images are allocated to a dataset that we used for pretraining.
- Spatially-varying label smoothing: each nucleus is assigned a label of 1 at its centroid, then each neighbouring pixel within the same nucleus would have a decaying probability of being part of it, with the borders having a probability of 0.5.

***Data augmentation***

- No additional augmentation methods are employed.

***Segmentation task method***

- Tissue: a multi-fold training of an encoder-decoder network. The encoder, pretrained on Imagenet, is a Mixed Vision-Transformer, and the decoder a Feature-Pyramid Network.
- Nuclei: same as for tissue segmentation, each category of nuclei is segmented in a semantic segmentation manner.

***Architecture***

- The encoder, pretrained on ImageNet, is a Mixed Vision-Transformer.
- The decoder is a Feature-Pyramid Network.

***Post-processing***

- No additional post-processing methods are employed.

***Results***

- Track1: Average F1 score: 0.71. Average micro dice score: 0.62. Final rank: 5.
- Track2: Average F1 score: 0.48. Average micro dice score: 0.62. Final rank: 3.

***Processing in validation cohort***

No specific changes were implemented for validation of the algorithm.

***Appendix A.5. Method 5: Biototem***

No description of the employed methodology was provided by the participants.

***Results***

- Track1: Average F1 score: 0.72. Average micro dice score: 0.73. Final rank: 4.
- Track2: Average F1 score: 0.42. Average micro dice score: 0.73. Final rank: 5.

***Appendix A.6. Method 6: HITSZLab***

Team name: HITSZLab

Authors: Zijie Fang, Jingyun Chen, Guoshuai Xu, Yongbing Zhang

Affiliation: Tsinghua University(Shenzhen), Harbin Institute of Technology(Shenzhen)

***Introduction***

A three-stage mechanism to train a panoptic segmentation framework for histopathological images of melanoma. Specifically, Mask2Former is trained for tissue segmentation for both Track 1 and Track 2. HoverNext is trained for cell segmentation in Track 1 and another Mask2Former is trained for cell segmentation in Track 2.

***Pre-processing***

- The 5120×5120-shaped context RoIs are split into non-overlapping 1024×1024 image patches.
- A threshold based filtering strategy is applied to exclude patches whose background area exceeds 20% of the total area.

***Data augmentation***

- Tissue: the input image is resized to 640 × 640 with random cropping and scaling, followed by random flips, translation, scaling, rotation, and optical distortion, each with a probability of 0.5.
- Both sliding window and test-time augmentation (TTA) strategies are employed for inference. The step size in the sliding window strategy is set to 320, and TTA involves random scaling within the range [0.5, 0.75, 1.0, 1.25, 1.5, 1.75] and random flipping to improve prediction stability.
- Nuclei: intensity scaling, gaussian noise injection, flipping, and random rotation are applied.

***Segmentation task method***

***Tissue***

- Stage 1: a Mask2Former is trained on the images of the region of interest (RoI), which have been annotated with pixel-level masks.
- Stage 2: the model is then employed to generate pseudo-masks for unannotated regions within the context images of RoIs.
- Stage 3: another Mask2Former is trained from scratch with the original RoIs and the split images along with their pseudo-masks.

***Nuclei, Track 1***

- Stage 1: the 1024×1024 RoIs are divided into 512×512-shaped patches with a stride of 256, which are used to train HoverNext.
- Stage 2: a sliding window strategy with the same patch size of 512 and a stride of 256 is employed to generate pseudo masks.
- Stage 3: the 1024×1024 images are directly used as the input for training HoverNext.

***Nuclei, Track 2***

- The cell instance maps are transformed to semantic maps by treating overlapped cells as one cell.
- Mask2Former is trained with the same hyperparameters and settings as those in the tissue segmentation stage.

***Architecture***

- Both Mask2Formers utilize the base model of Swin Transformer (Liu et al., 2021) as the encoder.
- In stage 1, the encoder of Mask2Formers is initialized with weights pre-trained on ImageNet-22K. In stage 3, the initialization is based on ImageNet-1K.

***Post-processing (nuclei, Track 2)***

- If a cell is predicted to be in the background, it is dropped.
- If a cell is predicted to be inside epidermis tissues, it is classified as an epidermal nucleus.
- If a cell is inside the tissues of the blood vessel, it is classified as the endothelium.

***Results***

- Track1: Average F1 score: 0.67. Average micro dice score: 0.62. Final rank: 6.
- Track2: Average F1 score: 0.42. Average micro dice score: 0.62. Final rank: 4.

***Appendix A.7. Method 7: Aira Matrix***

No description of the employed methodology was provided by the participants.

***Results***

- Track1: Average F1 score: 0.69. Average micro dice score: 0.56. Final rank: 7.
- Track2: Average F1 score: 0.45. Average micro dice score: 0.56. Final rank: 8.

***Appendix A.8. Method 8: mlafarge***

No description of the employed methodology was provided by the participants.

***Results***

- Track1: Average F1 score: 0.69. Average micro dice score: 0.59. Final rank: 10.
- Track2: Average F1 score: 0.44. Average micro dice score: 0.59. Final rank: 7.

***Appendix A.9. Method 9: UME***

Team name: UME

Authors: Negar Shahamiri, Moritz Rempe, Lukas Heine, Jens Kleesiek, Fabian Hörst

Affiliation: University Hospital Essen (AöR), German Cancer Consortium

(DKTK, Partner site Essen), TU Dortmund University

***Introduction***

This method emphasizes delivering a deployable solution within a 24hour development timeframe, using out-of-the-box frameworks. The pipeline combines two models, namely CellViT^++^ for nuclei detection and nnU-Net for tissue segmentation.

***Pre-processing***

- Pre-trained the nnU-Net network on the NSCLC dataset (Kludt et al., 2024) using one fold (1600 training, 400 validation images), selecting the best checkpoint based on validation performance.

***Data augmentation***

- No additional augmentation methods are employed.

***Segmentation task method***

- Tissue: pretrained on NSCLC dataset and finetuned on PUMA dataset with nnU-Net model(Isensee et al., 2021). A custom trainer is implemented with masked cross-entropy loss to ignore empty areas. A single fold is employed rather than nnU-Net’s default model ensemble.
- Nuclei: only trained the lightweight cell classifiers of CellViT^++^ model for the nuclei taxonomies corresponding to track 1 and track 2, respectively.

***Architecture***

• nnU-Net with the same setup as the challenge baseline for tissue segmentation.

- CellViT^++^ model is used without any modifications compared to the original publication (Hörst et al., 2025).

**Post-processing**

• No additional post-processing methods are employed.

***Results***

- Track1: Average F1 score: 0.70. Average micro dice score: 0.61. Final rank: 9.
- Track2: Average F1 score: 0.39. Average micro dice score: 0.61. Final rank: 9.

***Appendix A.10. Method 10: SUDA-AIA-HeLab***

Team name: SUDA-AIA-HeLab

Authors: Tong Wang, HongLiang He, LiRui Qi, Wei Han, Hanbin Huang

Affiliation: Soochow University

***Introduction***

This method uses SMILE and nnU-Net as segmentation models for nuclei and tissue segmentation, respectively. A custom loss function is designed to mitigate the class imbalance problem.

***Pre-processing***

- Tissue: images where the proportion of class 1 and class 3 exceeded 80% are excluded. Then, to boost the sample size for classes 2, 4, and 5, data augmentation and replication are applied, resulting in a total of 299 training images.
- Nuclei: a sliding window strategy is implemented to divide the original 1024×1024 pixel regions of interest (ROIs) into smaller patches. Specifically, 256×256 pixel windows are moved with a stride of 128 pixels, resulting in 49 patches from each ROI. In total, 10,045 labeled training images are generated.

***Data augmentation***

- Tissue: random rotation and scaling, gaussian noise, contrast and brightness adjustments, random blurring, and low-resolution simulations are applied.
- Nuclei: random horizontal and vertical flip are applied.

***Segmentation task method***

- Tissue: nnU-Net model (Isensee et al., 2021). The input size of the model is set to [896, 768], ensuring that it can capture fine-grained features while maintaining computational efficiency.
- Nuclei: SMILE model (Pan et al., 2023). In the first phase, the decoder was frozen, and the model was trained for 20 epochs with a learning rate of $1e-4$. In the second phase, both the encoder and decoder were trained for 300 epochs.

***Architecture***

- nnU-Net with a ResEnc L architecture, which incorporates residual connections within the standard nnU-Net framework.
- A custom loss function is designed, the weights for the loss components are set as np: 1, hv: 1, tp: 1, where ”np,” ”hv,” and ”tp” correspond to different components of the loss function.
- An imbalance-aware loss is implemented, taking into account the class distribution in both the three-class and ten-class scenarios.

***Post-processing***

- No additional post-processing methods are employed.

***Results***

- Track1: Average F1 score: 0.67. Average micro dice score: 0.46. Final rank: 11.
- Track2: Average F1 score: 0.39. Average micro dice score: 0.46. Final rank: 12.

***Appendix A.11. Method 11: MedSegMasters***

Team name: MedSegMasters

Authors: Jian Yang, WeiHan Li

Affiliation: Zhejiang University

***Introduction***

This method builds upon Prompt-NucSeg (Shui et al., 2025) by incorporating the interactions between cells and tissues for nuclei segmentation, and enhances nnUNet (Isensee et al., 2021) by specifically addressing the sparse necrotic regions.

***Pre-processing***

- No additional pre-processing methods are employed.

***Data augmentation***

- No additional augmentation methods are employed.

***Segmentation task method***

- Tissue: nnU-Net model.
- Nuclei: a model is built upon PromptNucSeg by incorporating the interactions between cells and tissues.

***Architecture***

- nnU-Net model is enhanced by specifically addressing the sparse necrotic regions. To improve segmentation results, a DeepLab-ResNet (Chen et al., 2017) is combined to separately identify necrotic areas.
- An auxiliary loss and extract features from tissue masks to assist in cell classification.

***Post-processing***

- No additional post-processing methods are employed.

***Results***

- Track2: Average F1 score: 0.38. Average micro dice score: 0.55. Final rank: 10.

**References (supplements)**

Adams, D., 2026. rictoo/hoverlink-puma: HoverLink v1.0: PUMA Challenge Submission. https://doi.org/10.5281/zenodo.18568867

Bankhead, P., Loughrey, M.B., Fernández, J.A., Dombrowski, Y., McArt, D.G., Dunne, P.D., McQuaid, S., Gray, R.T., Murray, L.J., Coleman, H.G., James, J.A., Salto-Tellez, M., Hamilton, P.W., 2017. QuPath: Open source software for digital pathology image analysis. Sci Rep 7, 16878. https://doi.org/10.1038/s41598-017-17204-5

Baumann, E., Dislich, B., Rumberger, J.L., Nagtegaal, I.D., Martınez, M.R., Zlobec, I., 2024. HoVer-NeXt: A Fast Nuclei Segmentation and Classification Pipeline for Next Generation Histopathology. Medical Imaging with Deep Learning.

Blank, C.U., Haanen, J.B., Ribas, A., Schumacher, T.N., 2016. The “cancer immunogram.” Science 352, 658–660. https://doi.org/10.1126/science.aaf2834

Casparie, M., Tiebosch, A.T.M.G., Burger, G., Blauwgeers, H., van de Pol, A., van Krieken, J.H.J.M., Meijer, G.A., 2007. Pathology databanking and biobanking in The Netherlands, a central role for PALGA, the nationwide histopathology and cytopathology data network and archive. Cell Oncol 29, 19–24. https://doi.org/10.1155/2007/971816

Chen, L.-C., Papandreou, G., Kokkinos, I., Murphy, K., Yuille, A.L., 2017. DeepLab: Semantic Image Segmentation with Deep Convolutional Nets, Atrous Convolution, and Fully Connected CRFs. https://doi.org/10.48550/arXiv.1606.00915

Duin, I.A.J. van, Schuiveling, M., Maat, L.S. ter, Amsterdam, W.A.C. van, Berkmortel, F. van den, Boers-Sonderen, M., Groot, J.W.B. de, Hospers, G.A.P., Kapiteijn, E., Labots, M., Piersma, D., Schrader, A.M.R., Vreugdenhil, G., Westgeest, H., Veta, M., Blokx, W.A.M., Diest, P.J. van, Suijkerbuijk, K.P.M., 2024. Baseline tumor-infiltrating lymphocyte patterns and response to immune checkpoint inhibition in metastatic cutaneous melanoma. European Journal of Cancer 208. https://doi.org/10.1016/j.ejca.2024.114190

Eisenhauer, E.A., Therasse, P., Bogaerts, J., Schwartz, L.H., Sargent, D., Ford, R., Dancey, J., Arbuck, S., Gwyther, S., Mooney, M., Rubinstein, L., Shankar, L., Dodd, L., Kaplan, R., Lacombe, D., Verweij, J., 2009. New response evaluation criteria in solid tumours: Revised RECIST guideline (version 1.1). European Journal of Cancer, Response assessment in solid tumours (RECIST): Version 1.1 and supporting papers 45, 228–247. https://doi.org/10.1016/j.ejca.2008.10.026

Fa’ak, F., Coudray, N., Jour, G., Ibrahim, M., Illa-Bochaca, I., Qiu, S., Claudio Quiros, A., Yuan, K., Johnson, D.B., Rimm, D.L., Weber, J.S., Tsirigos, A., Osman, I., 2025. Artificial Intelligence Algorithm Predicts Response to Immune Checkpoint Inhibitors. Clin Cancer Res 31, 3526–3536. https://doi.org/10.1158/1078-0432.CCR-24-3720

Gershenwald, J.E., Scolyer, R.A., Hess, K.R., Sondak, V.K., Long, G.V., Ross, M.I., Lazar, A.J., Faries, M.B., Kirkwood, J.M., McArthur, G.A., Haydu, L.E., Eggermont, A.M.M., Flaherty, K.T., Balch, C.M., Thompson, J.F., for members of the American Joint Committee on Cancer Melanoma Expert Panel and the International Melanoma Database and Discovery Platform, 2017. Melanoma staging: Evidence-based changes in the American Joint Committee on Cancer eighth edition cancer staging manual. CA: A Cancer Journal for Clinicians 67, 472–492. https://doi.org/10.3322/caac.21409

Giraud-Sauveur, F., Blampey, Q., Benkirane, H., Marinello, A., Cournède, P.-H., Christodoulidis, S., 2025. STHELAR, a multi-tissue dataset linking spatial transcriptomics and histology for cell type annotation. https://doi.org/10.1101/2025.07.11.664123

He, K., Zhang, X., Ren, S., Sun, J., 2016. Deep Residual Learning for Image Recognition, in: 2016 IEEE Conference on Computer Vision and Pattern Recognition (CVPR). Presented at the 2016 IEEE Conference on Computer Vision and Pattern Recognition (CVPR), IEEE, Las Vegas, NV, USA, pp. 770–778. https://doi.org/10.1109/CVPR.2016.90

Hörst, F., Rempe, M., Becker, H., Heine, L., Keyl, J., Kleesiek, J., 2025. CellViT++: Energy-Efficient and Adaptive Cell Segmentation and Classification Using Foundation Models. https://doi.org/10.48550/arXiv.2501.05269

Isensee, F., Jaeger, P.F., Kohl, S.A.A., Petersen, J., Maier-Hein, K.H., 2021. nnU-Net: a self-configuring method for deep learning-based biomedical image segmentation. Nat Methods 18, 203–211. https://doi.org/10.1038/s41592-020-01008-z

Jiaqi-Lv/KongNet_Inference_Main: Main Github repository for KongNet Inference Code [WWW Document], n.d. URL https://github.com/Jiaqi-Lv/KongNet_Inference_Main (accessed 12.10.25).

Jochems, A., Schouwenburg, M.G., Leeneman, B., Franken, M.G., van den Eertwegh, A.J.M., Haanen, J.B.A.G., Gelderblom, H., Uyl-de Groot, C.A., Aarts, M.J.B., van den Berkmortel, F.W.P.J., Blokx, W.A.M., Cardous-Ubbink, M.C., Groenewegen, G., de Groot, J.W.B., Hospers, G.A.P., Kapiteijn, E., Koornstra, R.H., Kruit, W.H., Louwman, M.W., Piersma, D., van Rijn, R.S., Ten Tije, A.J., Vreugdenhil, G., Wouters, M.W.J.M., van der Hoeven, J.J.M., 2017. Dutch Melanoma Treatment Registry: Quality assurance in the care of patients with metastatic melanoma in the Netherlands. Eur J Cancer 72, 156–165. https://doi.org/10.1016/j.ejca.2016.11.021

Kleczek, P., Jaworek-Korjakowska, J., Gorgon, M., 2020. A novel method for tissue segmentation in high-resolution H&E-stained histopathological whole-slide images. Computerized Medical Imaging and Graphics 79, 101686. https://doi.org/10.1016/j.compmedimag.2019.101686

Kludt, C., Wang, Y., Ahmad, W., Bychkov, A., Fukuoka, J., Gaisa, N., Kühnel, M., Jonigk, D., Pryalukhin, A., Mairinger, F., Klein, F., Schultheis, A.M., Seper, A., Hulla, W., Brägelmann, J., Michels, S., Klein, S., Quaas, A., Büttner, R., Tolkach, Y., 2024. Next-generation lung cancer pathology: Development and validation of diagnostic and prognostic algorithms. Cell Rep Med 5, 101697. https://doi.org/10.1016/j.xcrm.2024.101697

Lee, S., Oh, J.W., Hwang, S., Shen, J., Park, S., Kim, H., Chae, Y.K., Lee, S.-H., Choi, Y.-L., Chung, J., Shin, J., Song, H., Valero Puche, A., Yoo, D., Lee, T., Oum, C., Kim, J., Ali, S.M., Ock, C.-Y., 2025. Deep learning–powered H&E whole-slide image analysis of endothelial cells to characterize tumor vascular environment and correlate treatment outcome to immunotherapy. J Clin Oncol 43, 2578–2578. https://doi.org/10.1200/JCO.2025.43.16_suppl.2578

Liu, S., Amgad, M., More, D., Rathore, M.A., Salgado, R., Cooper, L.A.D., 2024. A panoptic segmentation dataset and deep-learning approach for explainable scoring of tumor-infiltrating lymphocytes. NPJ Breast Cancer 10, 52. https://doi.org/10.1038/s41523-024-00663-1

Liu, Z., Lin, Y., Cao, Y., Hu, H., Wei, Y., Zhang, Z., Lin, S., Guo, B., 2021. Swin Transformer: Hierarchical Vision Transformer using Shifted Windows, in: 2021 IEEE/CVF International Conference on Computer Vision (ICCV). Presented at the 2021 IEEE/CVF International Conference on Computer Vision (ICCV), pp. 9992–10002. https://doi.org/10.1109/ICCV48922.2021.00986

Lv, J., Nasir, E., Xu, K., Jahanifar, M., Chohan, B., Raza, S. e A., 2025a. KongNet: A Multi-headed Deep Learning Model for Detection and Classification of Nuclei in Histopathology Images. https://doi.org/10.48550/arXiv.2510.23559

Lv, J., Nasir, E.S., Xu, K., Jahanifar, M., Chohan, B.S., Elhaminia, B., Raza, S.E.A., 2025b. KongNet: A Multi-headed Deep Learning Model for Detection and Classification of Nuclei in Histopathology Images. https://doi.org/10.48550/arXiv.2510.23559

Lv, J., Zhu, Y., Tenorio, C.G.C., Chohan, B.S., Eastwood, M., Raza, S.E.A., 2025c. Leveraging Pathology Foundation Models for Panoptic Segmentation of Melanoma in H&E Images. https://doi.org/10.48550/arXiv.2507.13974

Maier-Hein, L., Reinke, A., Godau, P., Tizabi, M.D., Buettner, F., Christodoulou, E., Glocker, B., Isensee, F., Kleesiek, J., Kozubek, M., Reyes, M., Riegler, M.A., Wiesenfarth, M., Kavur, A.E., Sudre, C.H., Baumgartner, M., Eisenmann, M., Heckmann-Nötzel, D., Rädsch, T., Acion, L., Antonelli, M., Arbel, T., Bakas, S., Benis, A., Blaschko, M.B., Cardoso, M.J., Cheplygina, V., Cimini, B.A., Collins, G.S., Farahani, K., Ferrer, L., Galdran, A., van Ginneken, B., Haase, R., Hashimoto, D.A., Hoffman, M.M., Huisman, M., Jannin, P., Kahn, C.E., Kainmueller, D., Kainz, B., Karargyris, A., Karthikesalingam, A., Kofler, F., Kopp-Schneider, A., Kreshuk, A., Kurc, T., Landman, B.A., Litjens, G., Madani, A., Maier-Hein, K., Martel, A.L., Mattson, P., Meijering, E., Menze, B., Moons, K.G.M., Müller, H., Nichyporuk, B., Nickel, F., Petersen, J., Rajpoot, N., Rieke, N., Saez-Rodriguez, J., Sánchez, C.I., Shetty, S., van Smeden, M., Summers, R.M., Taha, A.A., Tiulpin, A., Tsaftaris, S.A., Van Calster, B., Varoquaux, G., Jäger, P.F., 2024. Metrics reloaded: recommendations for image analysis validation. Nat Methods 21, 195–212. https://doi.org/10.1038/s41592-023-02151-z

Paige, NYC, USA and Microsoft Research, Cambridge, MA USA, n.d. paige-ai/Virchow2 · Hugging Face.

Pan, X., Cheng, J., Hou, F., Lan, R., Lu, C., Li, L., Feng, Z., Wang, H., Liang, C., Liu, Zhenbing, Chen, X., Han, C., Liu, Zaiyi, 2023. SMILE: Cost-sensitive multi-task learning for nuclear segmentation and classification with imbalanced annotations. Medical Image Analysis 88, 102867. https://doi.org/10.1016/j.media.2023.102867

Patil, N.S., Nabet, B.Y., Müller, S., Koeppen, H., Zou, W., Giltnane, J., Au-Yeung, A., Srivats, S., Cheng, J.H., Takahashi, C., de Almeida, P.E., Chitre, A.S., Grogan, J.L., Rangell, L., Jayakar, S., Peterson, M., Hsia, A.W., O’Gorman, W.E., Ballinger, M., Banchereau, R., Shames, D.S., 2022. Intratumoral plasma cells predict outcomes to PD-L1 blockade in non-small cell lung cancer. Cancer Cell 40, 289-300.e4. https://doi.org/10.1016/j.ccell.2022.02.002

Rahib, L., Wehner, M.R., Matrisian, L.M., Nead, K.T., 2021. Estimated Projection of US Cancer Incidence and Death to 2040. JAMA Netw Open 4, e214708. https://doi.org/10.1001/jamanetworkopen.2021.4708

Ronneberger, O., Fischer, P., Brox, T., 2015. U-Net: Convolutional Networks for Biomedical Image Segmentation, in: Navab, N., Hornegger, J., Wells, W.M., Frangi, A.F. (Eds.), Medical Image Computing and Computer-Assisted Intervention – MICCAI 2015. Springer International Publishing, Cham, pp. 234–241. https://doi.org/10.1007/978-3-319-24574-4_28

Roy, A.G., Navab, N., Wachinger, C., 2018. Concurrent spatial and channel ‘squeeze & excitation’ in fully convolutional networks: 21st International Conference on Medical Image Computing and Computer Assisted Intervention, MICCAI 2018. Medical Image Computing and Computer Assisted Intervention – MICCAI 2018 - 21st International Conference, 2018, Proceedings, Lecture Notes in Computer Science (including subseries Lecture Notes in Artificial Intelligence and Lecture Notes in Bioinformatics) 421–429. https://doi.org/10.1007/978-3-030-00928-1_48

Russakovsky, O., Deng, J., Su, H., Krause, J., Satheesh, S., Ma, S., Huang, Z., Karpathy, A., Khosla, A., Bernstein, M., Berg, A.C., Fei-Fei, L., 2015. ImageNet Large Scale Visual Recognition Challenge. https://doi.org/10.48550/arXiv.1409.0575

Schuiveling, M., 2024. Melanoma Histopathology Dataset with Tissue and Nuclei Annotations. https://doi.org/10.5281/zenodo.10940193

Schuiveling, M., Blokx, W.A.M., Breimer, G.E., Suijkerbuijk, K.P.M., Eek, D., Veta, M., 2024. A Novel Dataset for Nuclei Segmentation in Melanoma Histopathology.

Schuiveling, M., Duin, I.V., Maat, L.S.T., Weerd, J.V. der, Berkmortel, F.V. den, Burgers, F., Boers-Sonderen, M., Labots, M., Groot, J.W.D., Haanen, J.B. a. G., Hospers, G., Kapiteijn, E., Piersma, D., Simkens, L.H., Westgeest, H.M., Diest, P.V., Pluim, J., Suijkerbuijk, K., Blokx, W., Veta, M., 2025. 36P Interpretable histomorphological subtypes linked to ICI response in advanced melanoma using AI-assisted histopathology analysis. ESMO Real World Data and Digital Oncology 10. https://doi.org/10.1016/j.esmorw.2025.100235

Schuiveling, Mark, Liu, H., Eek, D., Breimer, G.E., Suijkerbuijk, K.P.M., Blokx, W.A.M., Veta, M., 2025a. A novel dataset for nuclei and tissue segmentation in melanoma with baseline nuclei segmentation and tissue segmentation benchmarks. GigaScience 14, giaf011. https://doi.org/10.1093/gigascience/giaf011

Schuiveling, Mark, van Duin, I.A.J., Ter Maat, L.S., van der Weerd, J.C., Verheijden, R.J., van den Berkmortel, F., Blank, C.U., Breimer, G.E., Burgers, F.H., Boers-Sonderen, M., van den Eertwegh, A.J.M., de Groot, J.W.B., Haanen, J.B.A.G., Hospers, G.A.P., Kapiteijn, E., Piersma, D., Vreugdenhil, G., Westgeest, H., Schrader, A.M.R., Pluim, J.P.W., van Diest, P.J., Veta, M., Suijkerbuijk, K.P.M., Blokx, W.A.M., 2025b. Artificial Intelligence-Detected Tumor-Infiltrating Lymphocytes and Outcomes in Anti-PD-1-Based Treated Melanoma. JAMA Oncol e254072. https://doi.org/10.1001/jamaoncol.2025.4072

Shen, J., Choi, Y.-L., Lee, T., Kim, H., Chae, Y.K., Dulken, B.W., Bogdan, S., Huang, M., Fisher, G.A., Park, S., Lee, S.-H., Hwang, J.-E., Chung, J.-H., Kim, L., Song, H., Pereira, S., Shin, S., Lim, Y., Ahn, C.H., Kim, Seulki, Oum, C., Kim, Sukjun, Park, G., Song, S., Jung, W., Kim, Seokhwi, Bang, Y.-J., Mok, T.S.K., Ali, S.M., Ock, C.-Y., 2024. Inflamed immune phenotype predicts favorable clinical outcomes of immune checkpoint inhibitor therapy across multiple cancer types. J Immunother Cancer 12, e008339. https://doi.org/10.1136/jitc-2023-008339

Shui, Z., Zhang, Y., Yao, K., Zhu, C., Zheng, S., Li, J., Li, H., Sun, Y., Guo, R., Yang, L., 2025. Unleashing the Power of Prompt-Driven Nucleus Instance Segmentation, in: Leonardis, A., Ricci, E., Roth, S., Russakovsky, O., Sattler, T., Varol, G. (Eds.), Computer Vision – ECCV 2024, Lecture Notes in Computer Science. Springer Nature Switzerland, Cham, pp. 288–304. https://doi.org/10.1007/978-3-031-73383-3_17

Suijkerbuijk, K.P.M., van Eijs, M.J.M., van Wijk, F., Eggermont, A.M.M., 2024. Clinical and translational attributes of immune-related adverse events. Nature Cancer 5, 557–571. https://doi.org/10.1038/s43018-024-00730-3

Tan, M., Le, Q.V., 2021. EfficientNetV2: Smaller Models and Faster Training. https://doi.org/10.48550/arXiv.2104.00298

Torbati, N., Meshcheryakova, A., Woitek, R., Hatamikia, S., Mechtcheriakova, D., Mahbod, A., 2026. NucFuseRank: Dataset Fusion and Performance Ranking for Nuclei Instance Segmentation. https://doi.org/10.48550/arXiv.2601.20104

Torbati, N., Meshcheryakova, A., Woitek, R., Hatamikia, S., Mechtcheriakova, D., Mahbod, A., 2025. A Multi-Stage Auto-Context Deep Learning Framework for Tissue and Nuclei Segmentation and Classification in H&E-Stained Histological Images of Advanced Melanoma. https://doi.org/10.48550/arXiv.2503.23958

Wolchok, J.D., Chiarion-Sileni, V., Gonzalez, R., Grob, J.-J., Rutkowski, P., Lao, C.D., Cowey, C.L., Schadendorf, D., Wagstaff, J., Dummer, R., Ferrucci, P.F., Smylie, M., Butler, M.O., Hill, A., Márquez-Rodas, I., Haanen, J.B.A.G., Guidoboni, M., Maio, M., Schöffski, P., Carlino, M.S., Lebbé, C., McArthur, G., Ascierto, P.A., Daniels, G.A., Long, G.V., Bas, T., Ritchings, C., Larkin, J., Hodi, F.S., 2022. Long-Term Outcomes With Nivolumab Plus Ipilimumab or Nivolumab Alone Versus Ipilimumab in Patients With Advanced Melanoma. J Clin Oncol 40, 127–137. https://doi.org/10.1200/JCO.21.02229

Xie, E., Wang, W., Yu, Z., Anandkumar, A., Alvarez, J.M., Luo, P., 2021. SegFormer: Simple and Efficient Design for Semantic Segmentation with Transformers. https://doi.org/10.48550/arXiv.2105.15203

Zhang, S., Sun, L., Zuo, J., Feng, D., 2024. Tumor associated neutrophils governs tumor progression through an IL-10/STAT3/PD-L1 feedback signaling loop in lung cancer. Translational Oncology 40, 101866. https://doi.org/10.1016/j.tranon.2023.101866

Zhou, Z., Rahman Siddiquee, M.M., Tajbakhsh, N., Liang, J., 2018. UNet++: A Nested U-Net Architecture for Medical Image Segmentation, in: Stoyanov, D., Taylor, Z., Carneiro, G., Syeda-Mahmood, T., Martel, A., Maier-Hein, L., Tavares, J.M.R.S., Bradley, A., Papa, J.P., Belagiannis, V., Nascimento, J.C., Lu, Z., Conjeti, S., Moradi, M., Greenspan, H., Madabhushi, A. (Eds.), Deep Learning in Medical Image Analysis and Multimodal Learning for Clinical Decision Support. Springer International Publishing, Cham, pp. 3–11. https://doi.org/10.1007/978-3-030-00889-5_1
